# Genetic Causes of Sudden Cardiac Arrest in the Community

**DOI:** 10.1101/2024.12.08.24318665

**Authors:** Evan P. Kransdorf, Marco Mathias, Kotoka Nakamura, Jonathan Tyrer, Paul D. Pharaoh, Harpriya Chugh, Kyndaron Reinier, Zeynep Akdemir, Eric Boerwinkle, Bing Yu, Sumeet S. Chugh

## Abstract

**Background:** Annually 300,000 Americans experience sudden cardiac arrest (SCA). Studies in referral SCA cohorts have observed rare variants in genes associated with arrhythmia and cardiomyopathy. We sought to: (1) establish the population prevalence of rare disease-causing variants in a set of candidate genes and (2) confirm the association of disease-causing variants in these genes with SCA in two prospective population-based studies.

**Methods:** SCA patients (n=3264) were accrued from the Oregon Sudden Unexpected Death Study and the PREdiction of Sudden death in mulTi-ethnic cOmmunities (PRESTO) study and compared to control patients (n=13713) from the Atherosclerosis Risk in Communities (ARIC) study. Whole genome sequencing was performed. Disease-causing (likely pathogenic or pathogenic) variants in candidate genes associated with arrhythmia/cardiomyopathy were identified using updated American College of Medical Genetics and Genomics criteria. Gene-collapsing case-control analysis was performed using the conditional logistic regression-sequence kernel association test.

**Results:** We identified 300 disease-causing variants, the majority of which were in cardiomyopathy genes (71%). There were 136 patients (4.2%) in the SCA group and 351 patients (2.6%) in the control group with one or more disease-causing variants (OR 1.66, 95% confidence interval 1.33-2.07, p<0.001). We identified 13 genes associated with an increased risk of SCA, nine associated with cardiomyopathy (*BAG3, DSC2, DSG2, FLNC, LMNA, MYBPC3, TNNI3, TNNT2, TTN*) and four with arrhythmia (*CACNA1C, CASQ2, KCNH2, KCNQ1*).

**Conclusions:** Disease-causing variants in cardiomyopathy genes were the predominant genetic cause of SCA. These findings inform which genes to include in genetic screening for SCA.

## Introduction

Each year 300,000 Americans will suffer a sudden cardiac arrest (SCA) with a mortality rate of 90% ^1^. Without the use of genetic testing, the cause of SCA remains unidentified in approximately 30% of cases where an autopsy is performed ^2^. Studies using gene-panel or exome sequencing have identified rare, heterozygous variants in genes associated with Mendelian causes of arrhythmia and cardiomyopathy in 10-20% of patients with SCA ^3–7^.

Previous studies examining the genetic basis of SCA may have been affected by several biases. First, these studies have primarily studied individuals of European ancestry that have been referred to specialized cardiogenetic centers ^6,8^. Other studies have been performed using participants in longitudinal cohort studies, where SCA was determined retrospectively and ischemic versus nonischemic etiology could not be established ^3^. Lastly, previous genetic studies of victims of SCA have utilized the American College of Medical Genetics and Genomics (ACMG) 3.0 sequence variant interpretation criteria ^9^ and several updates have been made to these criteria by the National Institutes of Health (NIH) Clinical Genome Resource (ClinGen) consortium since that time ^10^.

We have evaluated and previously reported the epidemiology of the detailed SCA phenotype in two ongoing community-based studies: the Oregon Sudden Unexpected Death Study (SUDS, since 2002) ^11,12^ and the PREdiction of Sudden death in mulTi-ethnic cOmmunities (PRESTO, since 2015) study ^13^. We sought to: (1) establish the population prevalence of rare disease-causing variants in a set of candidate genes and (2) confirm the association of disease-causing variants in these genes with SCA in these two prospective population-based studies.

## Methods

### SCA and Control Samples

SCA patients were accrued from the Oregon Sudden Unexpected Death Study (SUDS), Multnomah County, Oregon (n=2512)^11,12^ and the PREdiction of Sudden death in mulTi-ethnic cOmmunities (PRESTO) study Ventura County, California (n=966)^13^ (Figure 1). Patients were excluded if DNA sequencing did not meet quality control criteria (n=148 from SUDS and 66 from PRESTO). Based on review of available medical records the cause of SCA was assigned as definite or probable ischemic, definite or probable nonischemic, or unassigned if no records were available. Methods used for classification have been published previously ^14^ and are detailed in the Supplementary Appendix.

**Figure 1:**
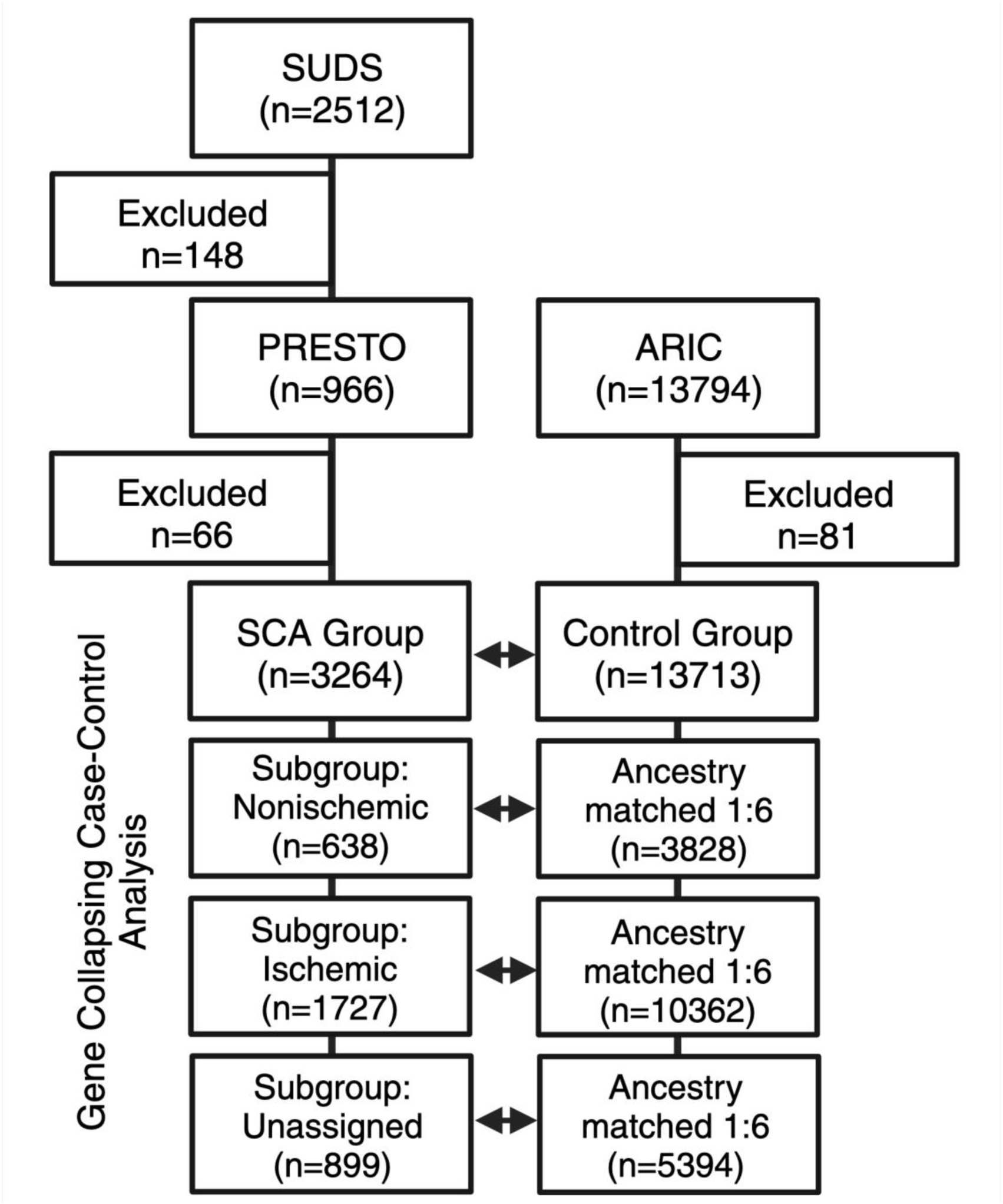
Design of the SCA and Control Groups. CONSORT diagram showing composition of the SCA and control groups. ARIC: Atherosclerosis Risk in Communities; PRESTO: PREdiction of Sudden death in mulTi-ethnic cOmmunities; SUDS: Oregon Sudden Unexpected Death Study.

Control patients (n=13713) were obtained from the Atherosclerosis Risk in Communities (ARIC) study and were persons without cardiac disease and aged 45 at baseline ^15^. Patients were excluded if DNA sequencing did not meet quality control criteria (n=81).

### Whole Genome Sequencing and Variant Calling

Whole genome sequencing was performed at Baylor College of Medicine Human Genome Sequencing Center. DNA samples were constructed into libraries according to the manufacturer’s protocol. All samples underwent paired-end sequencing using NovoSeq instruments (Illumina, San Diego, CA). Variant calling was performed jointly for the SCA and control groups using the DRAGEN 4.0 pipeline.

### Gene Selection and Filtering

We utilized the NIH Clinical Genome Resource (ClinGen) Gene Curation Expert Panels (GCEP) to identify genes associated with each cardiac phenotype. We specifically utilized the GCEPs for: (1) arrhythmic right ventricular cardiomyopathy^16^, (2) Brugada syndrome^17^, (3) catecholaminergic polymorphic ventricular tachycardia and short QT syndrome ^18^, (4) DCM^19^, (5) hypertrophic cardiomyopathy^20^, and (6) long QT syndrome^21^. Genes classified by each GCEP as being definitively, strongly, moderately, or of limited association were included (Supplementary Table 1). The analysis was restricted to genes with autosomal dominant, semi-dominant and x-linked inheritance mechanisms. Variants were excluded if they were present in gnomAD exomes v4.1 or genomes 4.0 with total population frequency that was ≥ 0.1% ^22^ and if genotype quality was < 20.

### Variant Interpretation

Variants were assigned as benign/likely benign or likely pathogenic/pathogenic if the variant had that classification in ClinVar (version June 30, 2024)^23^ and a review status of “reviewed_by_expert_panel” or “criteria_provided,_multiple_submitters,_no_conflicts.” If no applicable ClinVar classification was present, variant interpretation was performed using the ACMG 3.0 criteria ^9^ with updates by ClinGen. A full specification of our variant interpretation criteria can be found in the Supplementary Appendix. Variant effect was determined using Ensembl Variant Effect Predictor ^24^ on Matched Annotation from the NCBI and EMBL-EBI (MANE) transcripts^25^. Proband counts were taken from submissions to ClinVar (version July 2024) with only the following phenotypes: cardiomyopathy (including arrhythmic, arrhythmogenic, dilated, hypertrophic), Brugada syndrome, catecholaminergic polymorphic ventricular tachycardia, long QT syndrome, short QT syndrome, sudden cardiac arrest/death, and ventricular fibrillation/tachycardia. Final variant pathogenicity classification was based on the sum of points awarded from all criteria ^26^.

### Automated Variant Interpretation

We developed an automated algorithm termed Python Automatic Variant Interpretation (PAVI) to apply our ACMG criteria. We validated PAVI by comparing automated interpretation against manual interpretation on a dataset of 60 patients with dilated cardiomyopathy and demonstrated 100% agreement.

### Statistical Analysis

Genetic ancestry was determined using principal component analysis applied to 33,661 uncorrelated SNPs (pairwise r2 < 0.1) with minor allele frequency >0.05 using an in-house program ^27^. Four major ancestry groups – White, Black, Asian, and Other/Unknown – were estimated from the first three principal components, using the four vertices of a tetrahedron.

The proportions of each ancestry for each sample were determined by identifying the nearest point on the tetrahedron and applying barycentric coordinates. Genetic ancestry correlated well with self-reported race (Supplementary Figure 1 and Supplementary Table 2).

For the total SCA group, patients were matched to those in ARIC 1:4. For the SCA phenotype subgroups, patients were matched to those in ARIC 1:6 by the first six principal components of genetic ancestry using the R package “optmatch” ^28^. Logistic modeling was used to assess the association between rare variants in the total SCA group and SCA phenotype subgroups and SCA using the “b.collapse” function in EPACTS with the first four principal components of genetic ancestry as covariates ^29^. Gene-collapsing rare variant case-control analysis was performed using conditional logistic regression-sequence kernel association test as implemented in CLoMAT/EPACTS ^30^.

All analyses and data visualizations were carried out using R as implemented in R Studio with the R packages dplyr and ggplot2.

## Results

### SCA and Control Groups

We included 3264 SCA patients from the SUDS (n=2364 or 72%) and PRESTO (n=900 or 28%) studies and 13713 control patients from the ARIC study (Figure 1). The SCA and control patients exhibited significant epidemiologic differences (Table 1), with SCA patients being older (66.0 vs. 54.0 years, p<0.001), less frequently women (29.5% vs. 55.5%, p<0.001), and more frequently self-reporting as White (88.4% vs. 75.8%, p<0.001). Genomic ancestry closely matched to self-reported race (Supplementary Figure 1 and Supplementary Table 2). Cause of SCA was determined to be definite/probable nonischemic in 638 (20%), definite/probable ischemic in 1727 (53%), and unassigned in 899 patients (27%) in the SCA group.

**Table 1:**
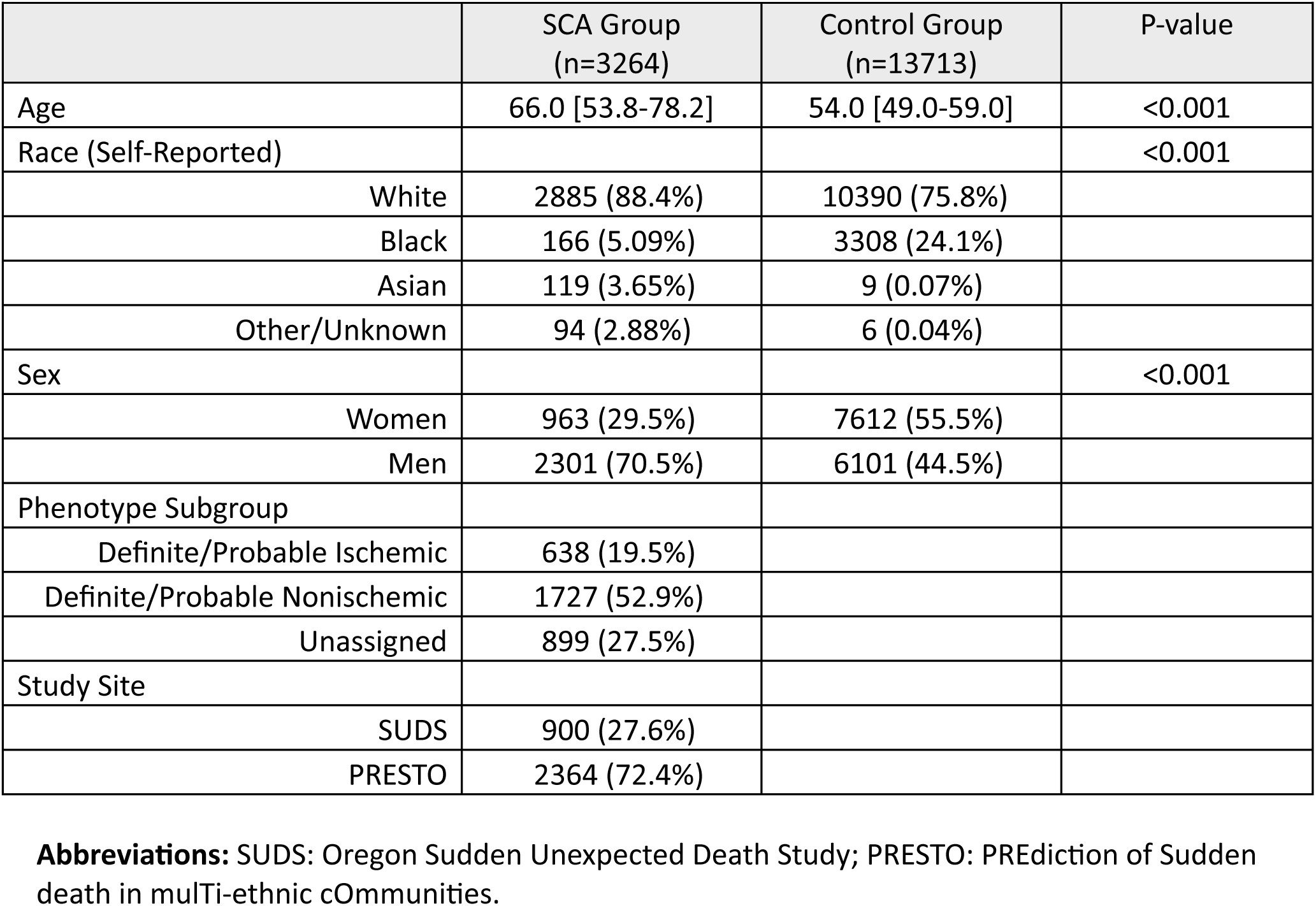
Characteristics of the SCA and Control Group.

### Gene Variants

We evaluated for disease-causing variants (likely pathogenic/pathogenic [LP/P] by ACMG sequence variant interpretation criteria) in 64 candidate genes previously associated with Mendelian arrhythmia and cardiomyopathy (Supplementary Table 1). We identified a total of 728,472 variants in these genes with population frequency <0.1% (Supplementary Table 2), of which 300 (0.04%) were classified as LP/P. We classified the remainder as benign/likely benign (5,903; 0.8%) or uncertain (722,269; 99.2%).

The 300 LP/P variants were present in 32 candidate genes (Supplementary Table 3); 136 were previously observed and classified in ClinVar as LP/P with high confidence and 164 were novel. Variant consequence (Supplementary Table 5) was frameshift/stop-gain in 166 (55%), missense in 92 (31%), canonical splice site in 31 (10%), and other consequence in 11 (4%). Frameshift/stop-gain variants were more frequent in the novel variant group (100/164 vs. 66/136; χ^2^ p-value=0.041) but there was no difference in the frequency of missense variants between the groups (49/164 vs. 43/136; χ^2^ p-value=0.842).

The number of LP/P variants per gene varied between one for *EYA4*, *KCNJ2*, *MYH6*, *MYL3*, *NEXN*, *TCAP*, *TPM1*, and 80 for *TTN*. There was a total of 505 variant allele heterozygotes for these 300 LP/P variants; 136 in SCA patients and 369 in controls. No variant homozygotes were detected in SCA patients or controls. Individual LP/P variants were uncommon, with the two highest frequency variants in SCA patients present in four (0.12%) patients each (variant chr7_150958449_G_A in the *KCNH2* gene and chr11_47342734_C_T in the *MYBPC3* gene). The highest frequency variant in control patients was present in 14 (0.10%) patients (variant chr3_38550865_A_G in *SCN5A* gene). Disease-causing variants were more prevalent in cardiomyopathy genes as compared to arrhythmia genes for both the SCA (102 vs. 34 of 136, 75%) and control (257 vs. 112 of 369, 70%) groups.

### Variants per Patient

We identified 133 patients (4.1%) in the total SCA group harboring 136 LP/P variants: 133 patients with a single variant and three patients each harboring two variants. We identified 351 patients (2.6%) in the control group harboring 369 LP/P variants: 334 patients with a single variant, 16 patients each harboring two LP/P variants, and one patient harboring three LP/P variants. The frequency of patients with at least one LP/P variant was higher in the SCA than the control group (OR 1.66, 95% confidence interval [95% CI] 1.33-2.07, p=6.89 x 10^-6^). The frequency of patients with two or more LP/P variants was similar in the SCA (0.09%) and control groups (0.1%).

There were 37 patients (5.8%) with LP/P variants in the nonischemic SCA subgroup, 61 patients (3.5%) in the ischemic SCA subgroup, and 35 patients (3.9%) in the unassigned subgroup (Figure 2). The frequency of patients with at least one LP/P variant was significantly higher in each of the SCA subgroups than in the control group: nonischemic SCA subgroup (OR 2.31, 95% CI 1.53-3.48, p=2.71 x 10^-4^), ischemic SCA subgroup (OR 1.42, 95% 95% CI 1.05-1.92, p=0.023), and the unassigned SCA subgroup (OR 1.64, 95% CI 1.11-2.44, p=0.013). The differences in odds ratios between the subgroups were not significant (P-heterogeneity = 0.17)

**Figure 2:**
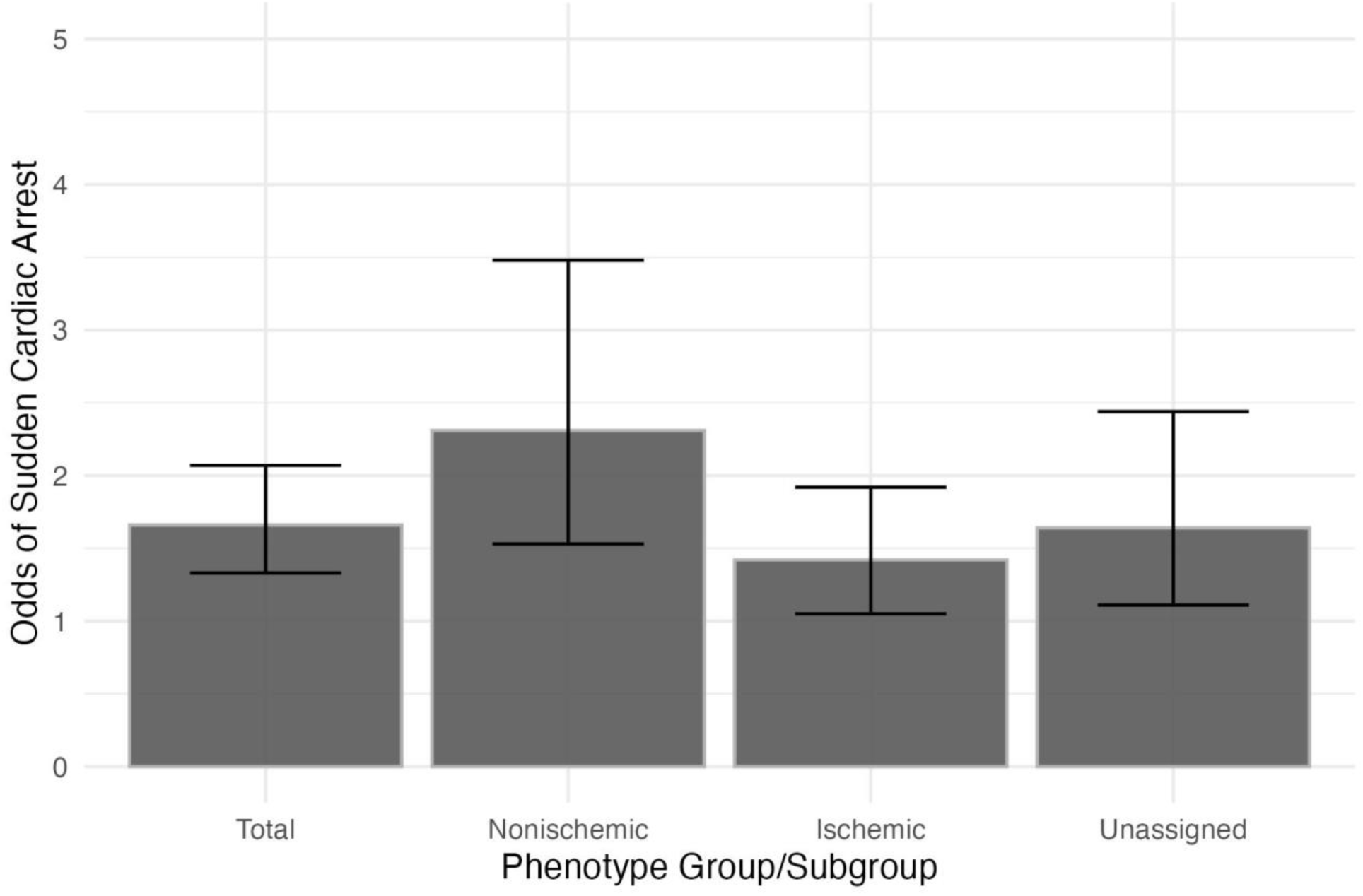
Odds of Sudden Cardiac Arrest With Disease-Causing Variants, by Group and Phenotype Subgroup. Barplot showing the odds of sudden cardiac arrest in patients harboring at least one disease-causing (likely pathogenic/pathogenic) variant in a candidate arrhythmia or cardiomyopathy gene for the total SCA cohort, nonischemic SCA subgroup, ischemic SCA subgroup, and unassigned SCA subgroup. Bars indicate 95% confidence intervals.

### Gene-Based Analysis

We then collapsed LP/P variants by gene and performed case-control analysis, while controlling for genomic ancestry, to assess the association between candidate genes harboring LP/P variants and the risk of SCA. For the total SCA group, one arrhythmia gene, *CACNA1C*, and two cardiomyopathy genes (*DSG2* and *LMNA)* were associated with a significantly increased risk of SCA (Table 2 and Supplementary Table 6). In the nonischemic SCA three arrhythmia genes (*CASQ2*, *KCNH2*, and *KCNQ1)* and seven cardiomyopathy genes (*BAG3*, *DSC2*, *FLNC*, *LMNA*, *MYBPC3, TNNI3*, and *TTN*) were associated with a significantly increased risk of SCA (Table 2 and Supplementary Table 7). In the ischemic SCA subgroup one arrhythmia gene, *CACNA1C,* and four cardiomyopathy genes (*DSG2*, *FLNC*, *LMNA,* and *TNNT2*) were associated with a significantly increased risk of SCA (Table 4 and Supplementary Table 8). Finally, in the unassigned SCA subgroup two cardiomyopathy genes, *DSG2* and *MYBPC3*, was associated with a significantly increased risk of SCA (Table 4 and Supplementary Table 9).

**Table 2:**
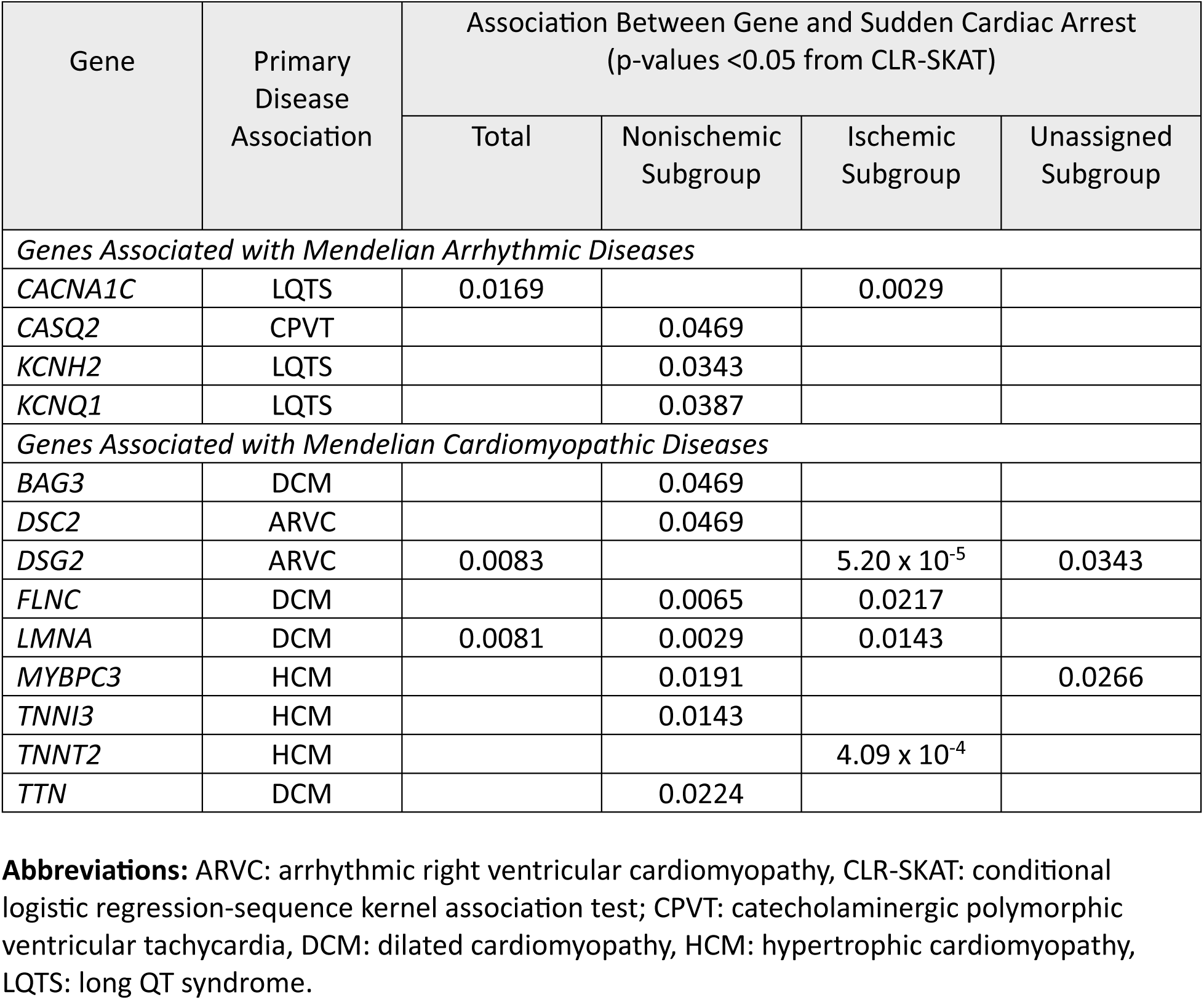
Genes Associated with Sudden Cardiac Arrest in Gene Collapsing Case-Control Analysis.

| Gene | Primary Disease Association | Association Between Gene and Sudden Cardiac Arrest (p-values <0.05 from CLR-SKAT) |  |  |  |
| --- | --- | --- | --- | --- | --- |
|  |  | Total | Nonischemic Subgroup | Ischemic Subgroup | Unassigned Subgroup |
| Genes Associated with Mendelian Arrhythmic Diseases |  |  |  |  |  |
| CACNA1C | LQTS | 0.0169 |  | 0.0029 |  |
| CASQ2 | CPVT |  | 0.0469 |  |  |
| KCNH2 | LQTS |  | 0.0343 |  |  |
| KCNQ1 | LQTS |  | 0.0387 |  |  |
| Genes Associated with Mendelian Cardiomyopathic Diseases |  |  |  |  |  |
| BAG3 | DCM |  | 0.0469 |  |  |
| DSC2 | ARVC |  | 0.0469 |  |  |
| DSG2 | ARVC | 0.0083 |  | 5.20 x 10<sup>-5</sup> | 0.0343 |
| FLNC | DCM |  | 0.0065 | 0.0217 |  |
| LMNA | DCM | 0.0081 | 0.0029 | 0.0143 |  |
| MYBPC3 | HCM |  | 0.0191 |  | 0.0266 |
| TNNI3 | HCM |  | 0.0143 |  |  |
| TNNT2 | HCM |  |  | 4.09 x 10<sup>-4</sup> |  |
| TTN | DCM |  | 0.0224 |  |  |

**Abbreviations:** ARVC: arrhythmic right ventricular cardiomyopathy, CLR-SKAT: conditional logistic regression-sequence kernel association test; CPVT: catecholaminergic polymorphic ventricular tachycardia, DCM: dilated cardiomyopathy, HCM: hypertrophic cardiomyopathy, LQTS: long QT syndrome.

## Discussion

In this community-based study of 64 candidate genes associated with Mendelian arrhythmia and cardiomyopathy, we identified disease-causing variants in 4.2% of SCA patients which was significantly higher than 2.6% in the control group. In patients with nonischemic SCA, the percentage was higher at 5.8%. We identified 13 genes, four causing arrhythmia and nine causing cardiomyopathy, which were associated with a significantly increased risk of SCA in the total SCA group or its subgroups compared to the control group. It is notable that 76% of the disease-causing variants we identified in the SCA group and nine of 13 genes that increased the risk of SCA are associated with cardiomyopathy, suggesting that cardiomyopathy is a greater contributor to SCA in the general population than Mendelian arrhythmic diseases. Although several of the cardiomyopathy genes we observed to be associated with SCA are well established causes of arrhythmic cardiomyopathy, our results suggest that cardiomyopathies due to disease-causing variants in *MYBC3, TNNI3,* and *TNNT2* should also be classified as arrhythmic cardiomyopathies.

Our current results are consistent with our previous study of the genetics of SCA in young patients in the community^7^ which yielded a lower prevalence of disease-causing variants as compared to studies performed in referral populations, where the typical prevalence of disease-causing variants is approximately 10% to 20%^4,8^. We identified disease-causing variants in only 32 of the 64 candidate genes, and only nine of these genes were associated with a significantly increased risk of SCA. Thus, our results suggest that a limited cadre of genes are responsible for SCA in the general population.

Current guidelines recommend genetic testing for patients with SCA^31^ but at this time there are no consensus recommendations as to what genes should be tested. This is an important consideration since SCA can be the sequelae of several Mendelian cardiac diseases (arrhythmic and cardiomyopathic) as well as non-Mendelian diseases such atherosclerotic coronary artery disease. The data presented here inform which genes should be considered as essential components of a genetic testing panel for a victim of SCA or their family members.

Current guidelines for the management of HCM and DCM recommend a phenotype- and risk factor-based approach to risk stratification for SCA^32,33^, but recent findings suggest that phenotype and risk factors may not always be a reliable starting point for risk stratification for SCA ^34,35^. Indeed, disease-causing genetic variants may cause SCA without clinically apparent heart disease ^5^. Furthermore, several cardiomyopathy genes can be associated with more than one cardiomyopathic phenotype, *e.g. MYBPC3* ^36^, can be associated with both HCM and DCM.

Thus, our data highlight the potential relevance of a gene centered, rather than phenotype centered, approach to genetic testing in cardiac disease.

Given that only 50% of patients with SCA have symptoms prior to arrest, SCA can be the first manifestation of a Mendelian arrhythmic or cardiomyopathic disease ^37^. As such, primary screening for disease-causing variants associated with an increased risk of SCA holds promise as an approach to reduce the incidence of SCA. The data presented here could be particularly informative with regards to which genes should be included in genetic screening for SCA, thus positively affecting the effectiveness of primary genetic screening ^38^.

Consistent with previous epidemiologic studies, definitive/probable ischemia was the most common phenotype of SCA. We identified four genes associated with an increased risk of SCA in the ischemic SCA subgroup, suggesting that disease-causing variants can contribute to ischemic SCA in a small percentage of cases. The mechanistic basis for this contribution is unclear, but previous studies have identified an increased risk of ischemic SCA in families ^39,40^, suggesting that it may have a rare variant genetic basis. Further investigation is needed to verify this intersection of phenotype and genotype.

This study has several strengths. First, given the large number of SCA cases in our study, we have been able to determine gene-based risks of SCA. Next, the availability of phenotyping for the cause of SCA in our study has allowed us to associate specific genes with causes of SCA. Our approach was limited by a single assessment of cardiovascular disease at baseline in the control group, and thus we cannot exclude the possibility that cardiovascular disease developed in control patients harboring LP/P variants over time.

In conclusion, we have found that 4.2% of SCA patients in our community-based group harbored a disease-causing variant. Furthermore, nine of the 13 genes causing an increased risk of SCA are associated with cardiomyopathy. These findings have implications for improving the genetic approach to primary and secondary risk stratification for SCA.

## Data Availability

All data will become available per NIH policies

