## Supplemental Material for "Genetic Causes of Sudden Cardiac Arrest in the Community"

**Supplementary Material**

Criteria for Sudden Cardiac Arrest Phenotypes

Supplementary Table 1: Candidate Genes

Variant Interpretation Criteria

Supplementary Figure 1: Principal Components Analysis of Genetic Ancestry

Supplementary Table 2: Comparison of Genetic Ancestry to Self-Reported Race

Supplementary Table 3: Frequency of Variants in Candidate Genes by American College of Genetics and Genomics Classification

Supplementary Table 4: Variants in Candidate Genes Classified as Likely Pathogenic/Pathogenic

Supplementary Table 5: Comparison of Variant Consequences Between Known and Novel Variants

Supplementary Table 6: Case-Control Analysis for Genes Associated with SCA, Sudden Cardiac Arrest-Total Cohort vs. Control Cohort

Supplementary Table 7 Case-Control Analysis for Genes Associated with SCA, Sudden Cardiac Arrest-Nonischemic Subgroup Matched 1:6 to Control Cohort

Supplementary Table 8: Case-Control Analysis for Genes Associated with SCA, Sudden Cardiac Arrest-Ischemic Subgroup Matched 1:6 to Control Cohort

Supplementary Table 9: Case-Control Analysis for Genes Associated with SCA, Sudden Cardiac Arrest-Unassigned Subgroup Matched 1:6 to Control Cohort

References

**Criteria for Sudden Cardiac Arrest Phenotypes**

1. **Definite Ischemic**
   a. Diagnosis of CAD (angiography/autopsy) defined as ≥ 50% stenosis in left main coronary artery or ≥70% stenosis in other coronary arteries
   b. Revascularization (PCI/CABG)
   c. Ischemic cardiomyopathy
   d. Myocardial infarction
2. **Probable ischemic**
   a. Diagnosis of CAD (without angiography/revascularization/autopsy)
   b. Stenosis by angiogram/autopsy between 50% and 69%
   c. CAD from physician dictation
   d. Positive stress test
   e. Unstable angina
3. **Definite Non-ischemic**
   a. Diagnosis of non-ischemic cardiac disease
      i. HCM, DCM, ARVC
      ii. Cardiac amyloidosis
      iii. Cardiac sarcoidosis
      iv. LQT, SQTS, BrS, CPVT
      v. Hypertensive cardiomyopathy, alcohol-related cardiomyopathy, nonspecific cardiomyopathy
      vi. Congenital heart disease
      vii. Recent myocarditis
      viii. Untreated WPW
      ix. SADS, idiopathic VF
      x. Coronary artery dissection
      xi. Nonspecific myocardial fibrosis (replacement/Interstitial) by autopsy, MRI (Late gadolinium)
      xii. Heart failure / systolic dysfunction (EF < 50%) and negative for CAD (findings of stenosis <70% from angiogram or autopsy)

**and**

b. No CAD diagnosis, ischemic cardiomyopathy, or MI

4. **Probable non-ischemic**
   a. Diagnosis of non-ischemic cardiac disease
      i. Heart failure / systolic dysfunction (EF < 50%) and no documentation of CAD on angiogram/autopsy **or**
      ii. Aortic stenosis (moderate or greater, Treated/untreated) **or**
      iii. Mitral valve prolapse (mitral annular disjunction) **or**
      iv. Aortic regurgitation (moderate or greater) **or**
      v. Prearrest myocarditis **or**
      vi. Prearrest conduction disorder (2^nd^/3rd-degree AV-block) **or**
      vii. Prearrest VF/VT/SCA/ablation **or**
      viii. Prearrest ICD/pacemaker **or**
      ix. Coronary anomalies **or**
      x. RV dysfunction/hypertrophy **or**
      xi. Wall motion abnormality (hypokinesia, akinesia, dyskinesia) **or**
      xii. Atrial fibrillation **or**
      xiii. Pre-arrest LVH in echocardiogram/ECG

**and**

b. No CAD diagnosis, ischemic cardiomyopathy, or MI

5. **Unassigned**
   a. No ischemic or non-ischemic substrate

**Supplementary Table 1: Classification of 64 ClinGen Candidate Genes**

| Gene | Primary GCEP | Primary Classification | Secondary GCEP | Secondary Classification | Additional GCEP |
| --- | --- | --- | --- | --- | --- |
| ABCC9 | DCM | Limited |  |  |  |
| ACTC1 | HCM | Definitive | DCM | Moderate |  |
| ANKRD1 | DCM | Limited |  |  |  |
| BAG3 | DCM | Definitive |  |  |  |
| CACNA1C | LQTS | Moderate |  |  |  |
| CALM1 | LQTS | Definitive | CPVT | Moderate |  |
| CALM2 | LQTS | Definitive | CPVT | Moderate |  |
| CALM3 | LQTS | Definitive | CPVT | Moderate |  |
| CASQ2 | CPVT | Moderate |  |  |  |
| CAV3 | LQTS | Limited |  |  |  |
| CDH2 | ARVC | Limited |  |  |  |
| CSRP3 | DCM | Limited | HCM | Moderate |  |
| CTF1 | DCM | Limited |  |  |  |
| CTNNA3 | ARVC | Limited |  |  |  |
| DES | DCM | Definitive | ARVC | Moderate |  |
| DSC2 | ARVC | Definitive |  |  |  |
| DSG2 | ARVC | Definitive | DCM | Limited |  |
| DSP | ARVC | Definitive |  |  |  |
| DTNA | DCM | Limited |  |  |  |
| EYA4 | DCM | Limited |  |  |  |
| FLNC | HCM | Definitive | DCM | Definitive |  |
| ILK | DCM | Limited |  |  |  |
| JPH2 | DCM | Moderate |  |  |  |
| KCNE1 | LQTS | Limited |  |  |  |
| KCNH2 | LQTS | Definitive | SQTS | Definitive |  |
| KCNJ2 | LQTS | Limited | SQTS | Moderate |  |
| KCNQ1 | LQTS | Definitive | SQTS | Strong |  |
| LAMA4 | DCM | Limited |  |  |  |
| LAMP2 | HCM | Definitive |  |  |  |
| LDB3 | DCM | Limited |  |  |  |
| LMNA | DCM | Definitive | ARVC | Limited |  |
| MYBPC3 | ARVC | Limited | DCM | Limited |  |
| MYH6 | HCM | Limited | DCM | Limited |  |
| MYH7 | DCM | Definitive | ARVC | Limited |  |
| MYL2 | HCM | Definitive | DCM | Limited |  |
| MYL3 | HCM | Definitive | ARVC | Limited |  |
| MYPN | DCM | Limited |  |  |  |
| NEBL | DCM | Limited |  |  |  |
| NEXN | DCM | Moderate |  |  |  |
| NKX2-5 | DCM | Limited |  |  |  |
| OBSCN | DCM | Limited |  |  |  |
| PKP2 | ARVC | Definitive |  |  |  |
| PLN | HCM | Definitive | ARVC | Moderate |  |
| PRDM16 | DCM | Limited |  |  |  |
| PRKAG2 | HCM | Definitive |  |  |  |
| PSEN2 | DCM | Limited |  |  |  |
| RBM20 | DCM | Definitive |  |  |  |
| RYR2 | CPVT | Definitive |  |  |  |
| SCN5A | BrS | Definitive | DCM | Definitive | SEE NOTE |
| SGCD | DCM | Limited |  |  |  |
| SLC4A3 | SQTS | Moderate |  |  |  |
| TBX20 | DCM | Limited |  |  |  |
| TCAP | DCM | Limited |  |  |  |
| TGFB3 | ARVC | Limited |  |  |  |
| TJP1 | ARVC | Limited |  |  |  |
| TMEM43 | ARVC | Definitive |  |  |  |
| TNNC1 | DCM | Definitive |  |  |  |
| TNNI3 | HCM | Definitive | DCM | Moderate |  |
| TNNI3K | DCM | Limited |  |  |  |
| TNNT2 | DCM | Definitive | HCM | Definitive |  |
| TPM1 | HCM | Definitive | DCM | Moderate |  |
| TTN | DCM | Definitive | ARVC | Limited |  |
| TTR | HCM | Definitive |  |  |  |
| VCL | DCM | Moderate |  |  |  |

**Abbreviations:** ARVC: arrhythmic right ventricular cardiomyopathy, BrS: Brugada syndrome, CPVT: catecholaminergic polymorphic ventricular tachycardia, DCM: dilated cardiomyopathy, GCEP: ClinGen Gene Curation Expert Panel, HCM: hypertrophic cardiomyopathy, LQTS: long QT syndrome, SQTS: short QT syndrome.

**Note:** SCN5A is also associated with LQTS classified definitive and ARVC classified as limited.

**Variant Interpretation Criteria**

Very strong evidence of pathogenicity

• PVS1, Null variant (nonsense, frameshift, canonical +/−1 or 2 splice sites, initiation
codon, single or multi-exon deletion) in a definitive/strong gene ^1^

Strong evidence of pathogenicity

• PVS1_Strong, Null variant (nonsense, frameshift, canonical +/−1 or 2 splice sites, initiation
codon, single or multi-exon deletion) in a moderate gene (with a mouse phenotype) ^1^

• PS1, Same amino acid change as a previously established pathogenic variant
regardless of nucleotide change

• PS3, Well-established *in vitro* or *in vivo* functional studies supportive of a damaging effect on the gene or gene product

• PS4_Strong, Variant identified in ≥10 unrelated probands with consistent phenotypes ^2^

Moderate evidence of pathogenicity

• PVS1_Moderate, Null variant (nonsense, frameshift, canonical +/−1 or 2 splice sites, initiation
codon, single or multi-exon deletion) in a moderate gene ^1^

• PM1, Located in a mutational hot spot and/or critical and well-established functional domain (*e.g*. active site of an enzyme) without benign variation

• PM4, Protein length changes due to stop-loss/in-frame deletions/insertions in a non-repeat region

• PM5, Novel missense change at an amino acid residue where a different missense change determined to be pathogenic has been seen before

• PS4_Moderate, Variant identified in ≥6 probands with consistent phenotypes ^2,3^

Supporting evidence of pathogenicity

• PM2, Absent from gnomAD or at extremely low frequency (<=0.05%) in all gnomAD nonfounder populations ^2,4,5^

• PP3, Multiple lines of computational evidence support a deleterious effect on the gene or gene product ^6^

| Method | Benign (BP4) | | | | Pathogenic (PP3) | | | |
| --- | --- | --- | --- | --- | --- | --- | --- | --- |
|  | Very Strong | Strong | Moderate | Supporting | Supporting | Moderate | Strong | Very Strong |
| REVEL | ≤0.003 | (0.003, 0.016] | (0.016, 0.183] | (0.183, 0.290] | [0.644, 0.773) | [0.773, 0.932) | ≥0.932 | – |

• PS4_Supporting, Variant identified in ≥2 probands with consistent phenotypes ^2,3^

Strong evidence of benign impact

• BS1, Allele frequency is >0.05% in any gnomAD nonfounder population ^2,4^

• BS3, Well-established *in vitro* or *in vivo* functional studies shows no damaging effect on protein function or splicing

Supporting evidence of benign impact

• BP1, Missense variant in a gene for which primarily truncating variants are known to cause disease

• BP3, In-frame deletions/insertions in a repetitive region without a known function

• BP4, Multiple lines of computational evidence suggest no impact on gene or gene product (conservation, evolutionary, splicing impact, etc) ^6^

| Method | Benign (BP4) | | | | Pathogenic (PP3) | | | |
| --- | --- | --- | --- | --- | --- | --- | --- | --- |
|  | Very Strong | Strong | Moderate | Supporting | Supporting | Moderate | Strong | Very Strong |
| REVEL | ≤0.003 | (0.003, 0.016] | (0.016, 0.183] | (0.183, 0.290] | [0.644, 0.773) | [0.773, 0.932) | ≥0.932 | – |

Point system ^7^

| Strength | Pathogenic | Benign |
| --- | --- | --- |
| Indeterminate | 0 | 0 |
| Supporting | 1 | −1 |
| Moderate | 2 | −2 |
| Strong | 4 | −4 |
| Very Strong | 8 | −8 |

| Classification | Point ranges |
| --- | --- |
| Pathogenic | ≥ 10 |
| Likely Pathogenic | 6 – 9 |
| Uncertain | 0 – 5 |
| Likely Benign | −1 – −6 |
| Benign | ≤ −7 |

**Supplementary Figure 1: Principal Components Analysis of Genetic Ancestry**

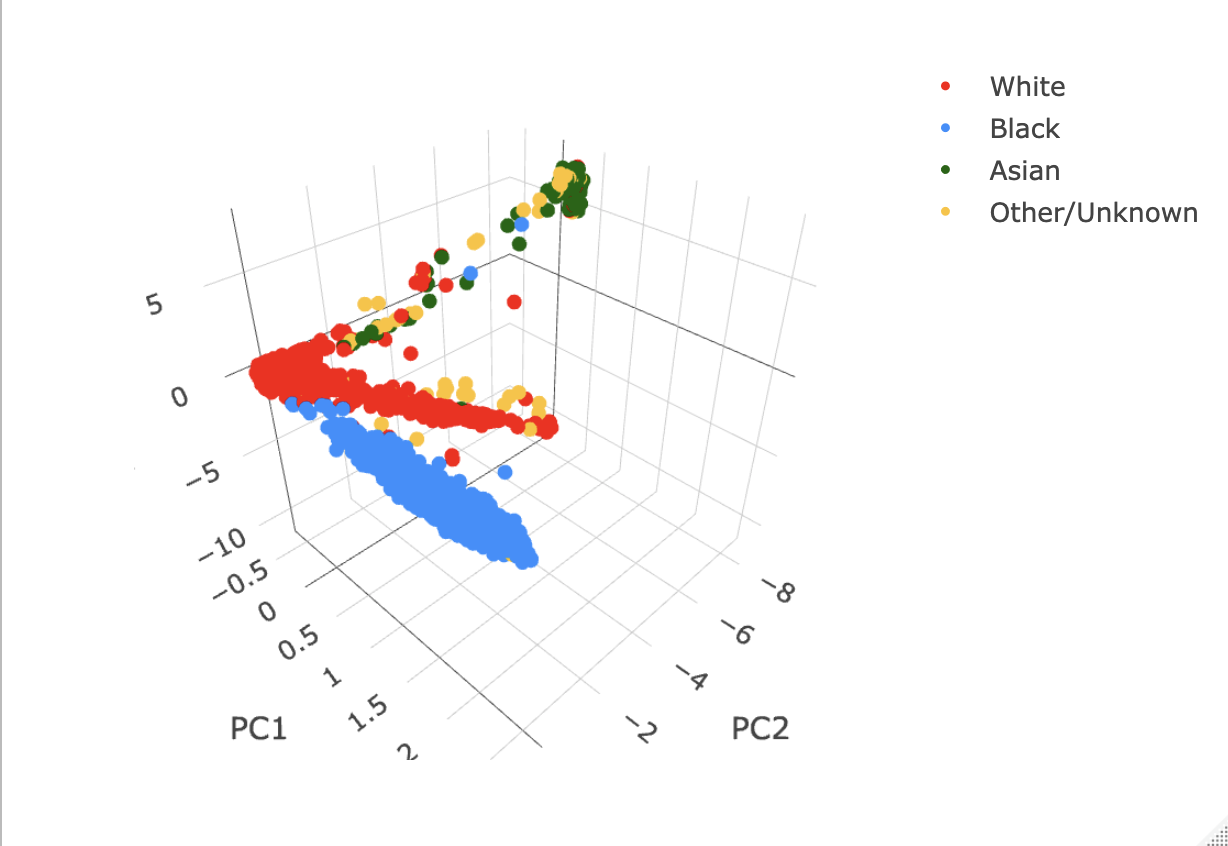

**Supplementary Figure 1:** Plot of principal component 1 (PC1), principal component 2 (PC2), and principal component 3 (PC3) of genetic ancestry as performed using OncoArray markers in the study cohort. Self-reported race is labeled by colors as per the legend.

**Supplementary Table 2: Comparison of Genetic Ancestry to Self-Reported Race**

| Genetic Ancestry | Self-Reported Race | | | | |
| --- | --- | --- | --- | --- | --- |
|  | Asian | Black | White | Other/Unknown | Total |
| African | 1  0.0% | 2265  98.8% | 9  0.4% | 17  0.7% | 2292  13.3% |
| Asian | 106  65.4% | 0  0.0% | 22  13.6% | 34  21.0% | 162  0.9% |
| European | 1  0.0% | 3  0.0% | 12712  98.1% | 244  1.9% | 12960  75.0% |
| Other/Mixed | 20  1.1% | 1206  64.9% | 532  28.6% | 100  5.4% | 1858  10.8% |
| Total | 128 | 3474 | 13275 | 62 | 17272 |

**Supplementary Table 3: Frequency of Variants in Candidate Genes by American College of Genetics and Genomics Classification**

|  |  | ACMG Classification | | |
| --- | --- | --- | --- | --- |
| Gene | Total  Variants | Likely Benign/  Benign | Uncertain Significance | Likely Pathogenic/  Pathogenic |
| ABCC9 | 11429 | 76 | 11353 | 0 |
| ACTC1 | 1125 | 28 | 1093 | 4 |
| ANKRD1 | 1596 | 29 | 1567 | 0 |
| BAG3 | 3957 | 49 | 3905 | 3 |
| CACNA1C | 52782 | 207 | 52573 | 2 |
| CALM1 | 1877 | 16 | 1861 | 0 |
| CALM2 | 3139 | 6 | 3133 | 0 |
| CALM3 | 1594 | 17 | 1577 | 0 |
| CASQ2 | 6908 | 25 | 6879 | 4 |
| CAV3 | 1912 | 20 | 1892 | 0 |
| CDH2 | 18109 | 83 | 18026 | 0 |
| CSRP3 | 2487 | 15 | 2470 | 2 |
| CTF1 | 1218 | 15 | 1203 | 0 |
| CTNNA3 | 140610 | 85 | 140525 | 0 |
| DES | 1382 | 25 | 1351 | 6 |
| DSC2 | 3709 | 70 | 3633 | 6 |
| DSG2 | 4175 | 73 | 4092 | 10 |
| DSP | 4182 | 172 | 3990 | 20 |
| DTNA | 13634 | 86 | 13548 | 0 |
| EYA4 | 22578 | 30 | 22547 | 1 |
| FLNC | 3132 | 269 | 2853 | 10 |
| ILK | 1308 | 41 | 1267 | 0 |
| JPH2 | 8771 | 98 | 8673 | 0 |
| KCNE1 | 6214 | 21 | 6193 | 0 |
| KCNH2 | 3644 | 100 | 3534 | 10 |
| KCNJ2 | 1562 | 29 | 1532 | 1 |
| KCNQ1 | 37489 | 195 | 37274 | 20 |
| LAMA4 | 10781 | 178 | 10603 | 0 |
| LAMP2 | 2772 | 30 | 2742 | 0 |
| LDB3 | 6225 | 130 | 6095 | 0 |
| LMNA | 2723 | 48 | 2672 | 3 |
| MYBPC3 | 3085 | 172 | 2890 | 23 |
| MYH6 | 3083 | 168 | 2914 | 1 |
| MYH7 | 2381 | 168 | 2192 | 21 |
| MYL2 | 1811 | 21 | 1785 | 5 |
| MYL3 | 1058 | 17 | 1040 | 1 |
| MYPN | 8814 | 138 | 8676 | 0 |
| NEBL | 10095 | 164 | 9931 | 0 |
| NEXN | 5577 | 49 | 5527 | 1 |
| NKX2-5 | 1221 | 17 | 1204 | 0 |
| OBSCN | 14641 | 246 | 14395 | 0 |
| PKP2 | 9160 | 71 | 9075 | 14 |
| PLN | 1874 | 7 | 1867 | 0 |
| PRDM16 | 39850 | 250 | 39600 | 0 |
| PRKAG2 | 30132 | 64 | 30068 | 0 |
| PSEN2 | 2733 | 16 | 2717 | 0 |
| RBM20 | 15893 | 76 | 15812 | 5 |
| RYR2 | 66413 | 311 | 66096 | 6 |
| SCN5A | 8486 | 166 | 8296 | 24 |
| SGCD | 35248 | 19 | 35226 | 3 |
| SLC4A3 | 1942 | 67 | 1875 | 0 |
| TBX20 | 4243 | 26 | 4217 | 0 |
| TCAP | 930 | 14 | 915 | 1 |
| TGFB3 | 2505 | 51 | 2454 | 0 |
| TJP1 | 11014 | 24 | 10990 | 0 |
| TMEM43 | 2339 | 64 | 2275 | 0 |
| TNNC1 | 926 | 17 | 909 | 0 |
| TNNI3 | 1564 | 42 | 1518 | 4 |
| TNNI3K | 24435 | 46 | 24389 | 0 |
| TNNT2 | 2206 | 36 | 2164 | 6 |
| TPM1 | 2625 | 38 | 2586 | 1 |
| TTN | 18086 | 981 | 17025 | 80 |
| TTR | 1245 | 16 | 1227 | 2 |
| VCL | 9833 | 75 | 9758 | 0 |
| TOTAL | 728472 | 5903 | 722269 | 300 |

**Supplementary Table 4: Variants in Candidate Genes Classified as Likely Pathogenic/Pathogenic**

| **Variant^1^** | **Gene** | **Coding** | **Protein** | **Consequence** | **ClinVar** | **Class** |
| --- | --- | --- | --- | --- | --- | --- |
| chr1_77942756_A_G | NEXN | ENST00000334785 c.1955A>G | ENSP00000333938 p.Tyr652Cys | MIS | 0 | LP |
| chr1_115701344_A_G | CASQ2 | ENST00000261448 c.1097T>C | ENSP00000261448 p.Leu366Pro | MIS | 1 | LP |
| chr1_115705208_G_A | CASQ2 | ENST00000261448  c.923C>T | ENSP00000261448 p.Pro308Leu | MIS | 1 | P/LP |
| chr1_115740767_G_A | CASQ2 | ENST00000261448  c.381C>T | ENSP00000261448  p. Gly127%3D | OTH | 1 | P/LP |
| chr1_115768378_T_C | CASQ2 | ENST00000261448  c.164A>G | ENSP00000261448 p.Tyr55Cys | MIS | 1 | P/LP |
| chr1_156135925_C_T | LMNA | ENST00000368300  c.961C>T | ENSP00000357283 p.Arg321Ter | SG | 1 | P |
| chr1_156136110_C_T | LMNA | ENST00000368300 c.1146C>T | ENSP00000357283 p.Gly382%3D | OTH | 1 | P/LP |
| chr1_156136952_G_A | LMNA | ENST00000368300 c.1412G>A | ENSP00000357283 p.Arg471His | MIS | 1 | P/LP |
| chr1_201362395_C_T | TNNT2 | ENST00000656932  c.601-1G>A | NA | SPL | 0 | P |
| chr1_201364335_CG_C | TNNT2 | ENST00000656932  c.451del | ENSP00000499593 p.Arg151GlyfsTer41 | FS | 0 | P |
| chr1_201364351_CG_C | TNNT2 | ENST00000656932  c.435del | ENSP00000499593 p.Glu146SerfsTer46 | FS | 0 | LP |
| chr1_201365617_T_G | TNNT2 | ENST00000656932  c.287A>C | ENSP00000499593 p.Asp96Ala | MIS | 1 | P/LP |
| chr1_201365622_T_TC | TNNT2 | ENST00000656932 c.281_282insG | ENSP00000499593 p.Val95SerfsTer4 | FS | 0 | P |
| chr1_201368162_C_A | TNNT2 | ENST00000656932  c.163G>T | ENSP00000499593 p.Glu55Ter | SG | 0 | P |
| chr1_237441428_T_A | RYR2 | ENST00000366574 c.1115T>A | ENSP00000355533 p.Leu372His | MIS | 0 | LP |
| chr1_237496525_T_G | RYR2 | ENST00000366574 c.1976T>G | ENSP00000355533 p.Ile659Ser | MIS | 0 | LP |
| chr1_237506807_A_G | RYR2 | ENST00000366574 c.2711A>G | ENSP00000355533 p.Tyr904Cys | MIS | 0 | LP |
| chr1_237614149_A_G | RYR2 | ENST00000366574 c.5021A>G | ENSP00000355533 p.His1674Arg | MIS | 0 | LP |
| chr1_237648612_C_T | RYR2 | ENST00000366574 c.7511C>T | ENSP00000355533 p.Thr2504Met | MIS | 0 | LP |
| chr1_237708988_CA_C | RYR2 | ENST00000366574 c.10033del | ENSP00000355533 p.Arg3345GlyfsTer12 | FS | 0 | LP |
| chr2_178527098_GT_G | TTN | ENST00000589042 c.107889del | ENSP00000467141 p.Lys35963AsnfsTer9 | FS | 1 | P |
| chr2_178527170_G_A | TTN | ENST00000589042 c.107818C>T | ENSP00000467141 p.Gln35940Ter | SG | 0 | LP |
| chr2_178527497_TA_T | TTN | ENST00000589042 c.107628del | ENSP00000467141 p.Asn35876LysfsTer2 | FS | 0 | LP |
| chr2_178528273_C_T | TTN | ENST00000589042 c.107377+1G>A | NA | SPL | 0 | P |
| chr2_178528629_GC_G | TTN | ENST00000589042 c.107121del | ENSP00000467141 p.Gln35707HisfsTer44 | FS | 0 | LP |
| chr2_178530418_GGT_G | TTN | ENST00000589042 c.106195_106196del | ENSP00000467141 p.Thr35399HisfsTer2 | FS | 0 | LP |
| chr2_178530823_T_TA | TTN | ENST00000589042 c.105791_105792insT | ENSP00000467141 p.Ser35265IlefsTer34 | FS | 1 | LP |
| chr2_178531355_GTC_G | TTN | ENST00000589042 c.105258_105259del | ENSP00000467141 p.Met35086IlefsTer11 | FS | 0 | LP |
| chr2_178531423_AAC_A | TTN | ENST00000589042 c.105190_105191del | ENSP00000467141 p.Val35064PhefsTer4 | FS | 1 | LP |
| chr2_178532215_TC_T | TTN | ENST00000589042 c.104399del | ENSP00000467141 p.Arg34800LysfsTer10 | FS | 0 | P |
| chr2_178532523_G_A | TTN | ENST00000589042 c.104092C>T | ENSP00000467141 p.Arg34698Ter | SG | 0 | P |
| chr2_178532669_C_CCCCAA | TTN | ENST00000589042 c.103945_103946insTTGGG | ENSP00000467141 p.Arg34649LeufsTer25 | FS | 0 | LP |
| chr2_178532670_GGT_G | TTN | ENST00000589042 c.103943_103944del | ENSP00000467141 p.Tyr34648SerfsTer3 | FS | 0 | LP |
| chr2_178533254_TC_T | TTN | ENST00000589042 c.103360del | ENSP00000467141 p.Glu34454AsnfsTer3 | FS | 0 | P |
| chr2_178533442_T_TG | TTN | ENST00000589042 c.103172_103173insC | ENSP00000467141 p.Pro34392ThrfsTer17 | FS | 0 | LP |
| chr2_178534383_C_A | TTN | ENST00000589042 c.102232G>T | ENSP00000467141 p.Glu34078Ter | SG | 0 | LP |
| chr2_178534887_C_A | TTN | ENST00000589042 c.101728G>T | ENSP00000467141 p.Glu33910Ter | SG | 1 | LP |
| chr2_178535516_T_TA | TTN | ENST00000589042 c.101098_101099insT | ENSP00000467141 p.Asp33700ValfsTer13 | FS | 1 | LP |
| chr2_178536971_C_CTA | TTN | ENST00000589042 c.100137_100138insTA | ENSP00000467141 p.Val33380Ter | FS | 0 | LP |
| chr2_178537800_CTG_C | TTN | ENST00000589042 c.99405_99406del | ENSP00000467141 p.Tyr33135Ter | SG | 0 | LP |
| chr2_178538834_GT_G | TTN | ENST00000589042 c.98994del | ENSP00000467141 p.Lys32998AsnfsTer63 | FS | 0 | P |
| chr2_178540194_G_A | TTN | ENST00000589042 c.97972C>T | ENSP00000467141 p.Arg32658Ter | SG | 1 | LP |
| chr2_178542723_G_GTA | TTN | ENST00000589042 c.97130_97131insTA | ENSP00000467141 p.Leu32379HisfsTer4 | FS | 1 | P/LP |
| chr2_178542739_C_CT | TTN | ENST00000589042 c.97114_97115insA | ENSP00000467141 p.Arg32372LysfsTer9 | FS | 1 | LP |
| chr2_178543549_CG_C | TTN | ENST00000589042 c.96423del | ENSP00000467141 p.Ile32141MetfsTer9 | FS | 0 | LP |
| chr2_178547445_G_GCT | TTN | ENST00000589042 c.94180_94181insAG | ENSP00000467141 p.Pro31394GlnfsTer3 | FS | 0 | LP |
| chr2_178547446_G_GCTAGA | TTN | ENST00000589042 c.94179_94180insTCTAG | ENSP00000467141 p.Pro31394SerfsTer4 | FS | 0 | LP |
| chr2_178548460_G_A | TTN | ENST00000589042 c.93166C>T | ENSP00000467141 p.Arg31056Ter | SG | 1 | P/LP |
| chr2_178549642_CACGGAATT_C | TTN | ENST00000589042 c.92072_92079del | ENSP00000467141 p.Gln30691ArgfsTer5 | FS | 0 | LP |
| chr2_178549728_AG_A | TTN | ENST00000589042 c.91993del | ENSP00000467141 p.Ala30666ProfsTer4 | FS | 0 | LP |
| chr2_178550122_A_AT | TTN | ENST00000589042 c.91715_91716insA | ENSP00000467141 p.Asn30572LysfsTer16 | FS | 0 | P |
| chr2_178551001_CTG_C | TTN | ENST00000589042 c.91528_91529del | ENSP00000467141 p.Gln30510ValfsTer10 | FS | 0 | LP |
| chr2_178552166_G_C | TTN | ENST00000589042 c.90734C>G | ENSP00000467141 p.Ser30245Ter | SG | 0 | LP |
| chr2_178552811_ACTT_ATT | TTN | ENST00000589042 c.90088del | ENSP00000467141 p.Val30030TyrfsTer10 | FS | 0 | LP |
| chr2_178552813_TTC_T | TTN | ENST00000589042 c.90085_90086del | ENSP00000467141 p.Glu30029SerfsTer7 | FS | 0 | LP |
| chr2_178554217_CTATT_C | TTN | ENST00000589042  c.88895-5_88895-2del | NA | SPL | 0 | LP |
| chr2_178559309_A_T | TTN | ENST00000589042 c.86821+2T>A | NA | SPL | 1 | P/LP |
| chr2_178559675_GTCATT_G | TTN | ENST00000589042 c.86452_86456del | ENSP00000467141 p.Asn28818LeufsTer51 | FS | 0 | LP |
| chr2_178560055_A_AT | TTN | ENST00000589042 c.86076_86077insA | ENSP00000467141 p.Ser28693IlefsTer2 | FS | 1 | P |
| chr2_178560869_T_TA | TTN | ENST00000589042 c.85262_85263insT | ENSP00000467141 p.Arg28422LysfsTer23 | FS | 0 | LP |
| chr2_178561117_GCTCA_G | TTN | ENST00000589042 c.85011_85014del | ENSP00000467141 p.Glu28338HisfsTer9 | FS | 0 | LP |
| chr2_178563058_TAGCACTTGCA_T | TTN | ENST00000589042 c.83064_83073del | ENSP00000467141 p.Ala27689LeufsTer31 | FS | 1 | P/LP |
| chr2_178565870_G_C | TTN | ENST00000589042 c.80262C>G | ENSP00000467141 p.Tyr26754Ter | SG | 0 | LP |
| chr2_178568469_AGAGTTACAATT_A | TTN | ENST00000589042 c.77652_77662del | ENSP00000467141 p.Glu25884AspfsTer3 | FS | 0 | LP |
| chr2_178568482_CGAT_CT | TTN | ENST00000589042 c.77648_77649del | ENSP00000467141 p.Ile25883ArgfsTer7 | FS | 0 | LP |
| chr2_178568485_TG_T | TTN | ENST00000589042 c.77646del | ENSP00000467141 p.Ile25883SerfsTer4 | FS | 0 | LP |
| chr2_178568947_T_A | TTN | ENST00000589042 c.77185A>T | ENSP00000467141 p.Lys25729Ter | SG | 1 | LP |
| chr2_178568981_CAG_C | TTN | ENST00000589042 c.77149_77150del | ENSP00000467141 p.Leu25717GlufsTer6 | FS | 0 | LP |
| chr2_178571425_G_A | TTN | ENST00000589042 c.74707C>T | ENSP00000467141 p.Gln24903Ter | SG | 0 | LP |
| chr2_178571440_C_A | TTN | ENST00000589042 c.74692G>T | ENSP00000467141 p.Glu24898Ter | SG | 0 | LP |
| chr2_178572593_A_AT | TTN | ENST00000589042 c.73538_73539insA | ENSP00000467141 p.Asn24513LysfsTer3 | FS | 0 | LP |
| chr2_178574276_C_CCCAGTATA | TTN | ENST00000589042 c.71855_71856insTATACTGG | ENSP00000467141 p.Gly23953IlefsTer23 | FS | 1 | LP |
| chr2_178575559_C_CTG | TTN | ENST00000589042 c.70572_70573insCA | ENSP00000467141 p.Glu23525GlnfsTer4 | FS | 0 | LP |
| chr2_178575970_G_A | TTN | ENST00000589042 c.70162C>T | ENSP00000467141 p.Arg23388Ter | SG | 1 | P/LP |
| chr2_178579559_A_G | TTN | ENST00000589042 c.67636+2T>C | NA | SPL | 1 | LP |
| chr2_178579775_CT_C | TTN | ENST00000589042 c.67421del | ENSP00000467141 p.Lys22474SerfsTer14 | FS | 1 | LP |
| chr2_178586613_CA_C | TTN | ENST00000589042 c.64287del | ENSP00000467141 p.Gly21430AlafsTer2 | FS | 1 | P/LP |
| chr2_178589849_G_A | TTN | ENST00000589042 c.61876C>T | ENSP00000467141 p.Arg20626Ter | SG | 1 | P/LP |
| chr2_178591043_T_TA | TTN | ENST00000589042 c.60681_60682insT | ENSP00000467141 p.Lys20228Ter | FS | 1 | LP |
| chr2_178592913_AC_A | TTN | ENST00000589042 c.59205del | ENSP00000467141 p.Glu19735AspfsTer24 | FS | 1 | P/LP |
| chr2_178594644_CA_C | TTN | ENST00000589042 c.57849del | ENSP00000467141 p.Val19284TyrfsTer18 | FS | 1 | LP |
| chr2_178598977_A_AT | TTN | ENST00000589042 c.56732_56733insA | ENSP00000467141 p.Asp18911GlufsTer25 | FS | 1 | LP |
| chr2_178600932_G_A | TTN | ENST00000589042 c.55972C>T | ENSP00000467141 p.Arg18658Ter | SG | 1 | LP |
| chr2_178601266_TATCGG_T | TTN | ENST00000589042 c.55726_55730del | ENSP00000467141 p.Pro18576IlefsTer4 | FS | 0 | LP |
| chr2_178601337_G_A | TTN | ENST00000589042 c.55660C>T | ENSP00000467141 p.Arg18554Ter | SG | 1 | LP |
| chr2_178601788_C_T | TTN | ENST00000589042  c.55303-1G>A | NA | SPL | 0 | P |
| chr2_178601915_C_T | TTN | ENST00000589042  c.55270-1G>A | NA | SPL | 0 | LP |
| chr2_178604986_C_T | TTN | ENST00000589042 c.54190+1G>A | NA | SPL | 1 | P/LP |
| chr2_178607520_AC_A | TTN | ENST00000589042 c.53167del | ENSP00000467141 p.Val17723LeufsTer7 | FS | 0 | LP |
| chr2_178608479_T_G | TTN | ENST00000589042  c.52406-2A>C | NA | SPL | 0 | LP |
| chr2_178610089_C_T | TTN | ENST00000589042 c.51436+1G>A | NA | SPL | 0 | P |
| chr2_178611568_G_GTATC | TTN | ENST00000589042 c.50660_50661insGATA | ENSP00000467141 p.Tyr16887Ter | SG | 1 | LP |
| chr2_178612115_G_A | TTN | ENST00000589042 c.50296C>T | ENSP00000467141 p.Arg16766Ter | SG | 1 | P/LP |
| chr2_178612355_G_A | TTN | ENST00000589042 c.50170C>T | ENSP00000467141 p.Arg16724Ter | SG | 1 | P/LP |
| chr2_178612527_AT_A | TTN | ENST00000589042 c.49997del | ENSP00000467141 p.Asn16666IlefsTer5 | FS | 0 | LP |
| chr2_178613020_TG_T | TTN | ENST00000589042 c.49700del | ENSP00000467141 p.Ser16567Ter | FS | 0 | LP |
| chr2_178613158_TA_T | TTN | ENST00000589042 c.49648+2del | NA | SPL | 1 | P |
| chr2_178649342_A_C | TTN | ENST00000589042  c.39974-11T>G | NA | SPL | 1 | P/LP |
| chr2_178739285_A_ACTTTT | TTN | ENST00000589042 c.13947_13948insAAAAG | ENSP00000467141 p.Phe4650LysfsTer14 | FS | 1 | LP |
| chr2_178740362_C_CT | TTN | ENST00000589042 c.12870_12871insA | ENSP00000467141 p.Val4291SerfsTer12 | FS | 1 | P/LP |
| chr2_219418687_GA_G | DES | ENST00000373960  c.226del | ENSP00000363071 p.Thr76ProfsTer22 | FS | 1 | P |
| chr2_219418869_T_A | DES | ENST00000373960  c.407T>A | ENSP00000363071 p.Leu136His | MIS | 0 | LP |
| chr2_219420575_CG_C | DES | ENST00000373960  c.817del | ENSP00000363071 p.Ala273ProfsTer31 | FS | 0 | LP |
| chr2_219420915_C_T | DES | ENST00000373960  c.985C>T | ENSP00000363071 p.Gln329Ter | SG | 0 | P |
| chr2_219421379_C_T | DES | ENST00000373960 c.1063C>T | ENSP00000363071 p.Arg355Ter | SG | 0 | LP |
| chr2_219425726_TCGAGA_T | DES | ENST00000373960 c.1354_1358del | ENSP00000363071 p.Glu452ThrfsTer8 | FS | 0 | LP |
| chr3_38550500_G_A | SCN5A | ENST00000423572 c.5869C>T | ENSP00000398266 p.Arg1957Ter | SG | 0 | LP |
| chr3_38550509_TCTCA_T | SCN5A | ENST00000423572 c.5856_5859del | ENSP00000398266 p.Ser1952ArgfsTer84 | FS | 0 | LP |
| chr3_38550634_C_T | SCN5A | ENST00000423572 c.5735G>A | ENSP00000398266 p.Arg1912His | MIS | 0 | LP |
| chr3_38550661_G_A | SCN5A | ENST00000423572 c.5708C>T | ENSP00000398266 p.Ser1903Leu | MIS | 0 | LP |
| chr3_38550865_A_G | SCN5A | ENST00000423572 c.5504T>C | ENSP00000398266 p.Ile1835Thr | MIS | 0 | LP |
| chr3_38550979_C_T | SCN5A | ENST00000423572 c.5390G>A | ENSP00000398266 p.Trp1797Ter | SG | 0 | LP |
| chr3_38551441_C_T | SCN5A | ENST00000423572 c.4928G>A | ENSP00000398266 p.Arg1643His | MIS | 1 | P |
| chr3_38551495_C_T | SCN5A | ENST00000423572 c.4874G>A | ENSP00000398266 p.Arg1625His | MIS | 0 | LP |
| chr3_38551519_GAGA_G | SCN5A | ENST00000423572 c.4847_4849del | ENSP00000398266 p.Phe1616del | OTH | 1 | P/LP |
| chr3_38554306_A_T | SCN5A | ENST00000423572 c.4783T>A | ENSP00000398266 p.Phe1595Ile | MIS | 0 | LP |
| chr3_38556481_AT_A | SCN5A | ENST00000423572 c.4393del | ENSP00000398266 p.Ile1465LeufsTer15 | FS | 0 | LP |
| chr3_38560179_C_T | SCN5A | ENST00000423572 c.4210G>A | ENSP00000398266 p.Val1404Met | MIS | 0 | LP |
| chr3_38562422_C_A | SCN5A | ENST00000423572 c.3953G>T | ENSP00000398266 p.Gly1318Val | MIS | 1 | P/LP |
| chr3_38580945_C_A | SCN5A | ENST00000423572 c.3214G>T | ENSP00000398266 p.Glu1072Ter | SG | 0 | LP |
| chr3_38585801_G_A | SCN5A | ENST00000423572 c.2677C>T | ENSP00000398266 p.Arg893Cys | MIS | 0 | LP |
| chr3_38585981_C_T | SCN5A | ENST00000423572 c.2497G>A | ENSP00000398266 p.Gly833Arg | MIS | 0 | LP |
| chr3_38603890_CT_C | SCN5A | ENST00000423572 c.1711del | ENSP00000398266 p.Ser571ValfsTer52 | FS | 0 | LP |
| chr3_38603940_G_GCT | SCN5A | ENST00000423572 c.1661_1662insAG | ENSP00000398266 p.Ser554ArgfsTer70 | FS | 0 | LP |
| chr3_38604031_G_GA | SCN5A | ENST00000423572 c.1570_1571insT | ENSP00000398266 p.Ser524PhefsTer17 | FS | 0 | LP |
| chr3_38606058_CG_C | SCN5A | ENST00000423572 c.1230del | ENSP00000398266 p.Val411TrpfsTer59 | FS | 0 | LP |
| chr3_38606682_C_T | SCN5A | ENST00000423572 c.1127G>A | ENSP00000398266 p.Arg376His | MIS | 1 | P/LP |
| chr3_38613773_G_A | SCN5A | ENST00000423572  c.673C>T | ENSP00000398266 p.Arg225Trp | MIS | 1 | P/LP |
| chr3_38613790_C_T | SCN5A | ENST00000423572  c.656G>A | ENSP00000398266 p.Arg219His | MIS | 0 | LP |
| chr3_38620843_G_A | SCN5A | ENST00000423572  c.611C>T | ENSP00000398266 p.Ala204Val | MIS | 0 | LP |
| chr3_46860813_G_T | MYL3 | ENST00000292327  c.170C>A | ENSP00000292327 p.Ala57Asp | MIS | 0 | LP |
| chr5_156508697_C_T | SGCD | ENST00000337851  c.289C>T | ENSP00000338343 p.Arg97Ter | SG | 1 | P/LP |
| chr5_156508703_G_A | SGCD | ENST00000337851 c.294+1G>A | NA | SPL | 1 | LP |
| chr5_156595042_C_T | SGCD | ENST00000337851  c.493C>T | ENSP00000338343 p.Arg165Ter | SG | 1 | P |
| chr6_7555737_AG_A | DSP | ENST00000379802  c.192del | ENSP00000369129 p.Arg64SerfsTer19 | FS | 0 | LP |
| chr6_7562718_CG_C | DSP | ENST00000379802  c.668del | ENSP00000369129 p.Gly223AlafsTer37 | FS | 0 | LP |
| chr6_7565521_G_A | DSP | ENST00000379802 c.939+1G>A | NA | SPL | 1 | P/LP |
| chr6_7570518_C_G | DSP | ENST00000379802 c.1656C>G | ENSP00000369129 p.Tyr552Ter | SG | 0 | LP |
| chr6_7574708_G_GTTAAAGGT | DSP | ENST00000379802 c.2353_2360dup | ENSP00000369129 p.Tyr787Ter | SG | 0 | LP |
| chr6_7574797_T_C | DSP | ENST00000379802 c.2436+2T>C | NA | SPL | 1 | P/LP |
| chr6_7575294_G_C | DSP | ENST00000379802  c.2437-1G>C | NA | SPL | 1 | LP |
| chr6_7576986_C_T | DSP | ENST00000379802 c.2821C>T | ENSP00000369129 p.Arg941Ter | SG | 1 | P/LP |
| chr6_7579323_C_T | DSP | ENST00000379802 c.3133C>T | ENSP00000369129 p.Arg1045Ter | SG | 1 | P/LP |
| chr6_7580055_C_T | DSP | ENST00000379802 c.3865C>T | ENSP00000369129 p.Gln1289Ter | SG | 1 | P/LP |
| chr6_7580671_CAAATGACCGG_C | DSP | ENST00000379802 c.4487_4496del | ENSP00000369129 p.Asp1496AlafsTer27 | FS | 0 | LP |
| chr6_7581258_GA_G | DSP | ENST00000379802 c.5070del | ENSP00000369129 p.Asp1691IlefsTer6 | FS | 0 | LP |
| chr6_7581402_C_T | DSP | ENST00000379802 c.5212C>T | ENSP00000369129 p.Arg1738Ter | SG | 1 | P |
| chr6_7581510_CAGAG_C | DSP | ENST00000379802 c.5327_5330del | ENSP00000369129 p.Glu1776GlyfsTer4 | FS | 1 | P/LP |
| chr6_7583604_TTTGA_T | DSP | ENST00000379802 c.6348_6351del | ENSP00000369129 p.Asp2117GlufsTer17 | FS | 1 | P/LP |
| chr6_7584385_G_C | DSP | ENST00000379802 c.7123G>C | ENSP00000369129 p.Gly2375Arg | MIS | 1 | LP |
| chr6_7585444_G_GA | DSP | ENST00000379802 c.8183dup | ENSP00000369129 p.Phe2729ValfsTer10 | FS | 0 | LP |
| chr6_7585701_A_AC | DSP | ENST00000379802 c.8442dup | ENSP00000369129 p.Ser2815GlnfsTer37 | FS | 0 | LP |
| chr6_7585776_C_CAG | DSP | ENST00000379802 c.8514_8515insAG | ENSP00000369129 p.Gly2840ProfsTer44 | FS | 0 | LP |
| chr6_7585777_TCC_T | DSP | ENST00000379802 c.8516_8517del | ENSP00000369129 p.Ser2839TrpfsTer12 | FS | 0 | LP |
| chr6_133462477_G_A | EYA4 | ENST00000355286  c.580G>A | ENSP00000347434 p.Asp194Asn | MIS | 1 | LP |
| chr7_128838037_T_G | FLNC | ENST00000325888 c.1020T>G | ENSP00000327145 p.Tyr340Ter | SG | 0 | LP |
| chr7_128841565_C_T | FLNC | ENST00000325888 c.2119C>T | ENSP00000327145 p.Gln707Ter | SG | 1 | P |
| chr7_128842339_TG_T | FLNC | ENST00000325888 c.2234del | ENSP00000327145 p.Gly745GlufsTer3 | FS | 1 | P |
| chr7_128843826_G_A | FLNC | ENST00000325888 c.2842G>A | ENSP00000327145 p.Gly948Arg | MIS | 0 | LP |
| chr7_128844656_A_G | FLNC | ENST00000325888  c.3193-2A>G | NA | SPL | 0 | P |
| chr7_128844769_C_T | FLNC | ENST00000325888 c.3304C>T | ENSP00000327145 p.Pro1102Ser | MIS | 0 | LP |
| chr7_128846089_G_A | FLNC | ENST00000325888 c.3890G>A | ENSP00000327145 p.Gly1297Glu | MIS | 0 | LP |
| chr7_128848974_T_TCGTCACAA | FLNC | ENST00000325888 c.4926_4927insACGTCACA | ENSP00000327145 p.Val1643ThrfsTer26 | FS | 1 | P/LP |
| chr7_128853012_C_CG | FLNC | ENST00000325888 c.6190dup | ENSP00000327145 p.Val2064GlyfsTer16 | FS | 1 | P/LP |
| chr7_128856636_CT_C | FLNC | ENST00000325888 c.7371del | ENSP00000327145 p.Glu2458SerfsTer71 | FS | 1 | P |
| chr7_150948445_C_CG | KCNH2 | ENST00000262186 c.2690_2691insC | ENSP00000262186 p.Lys897AsnfsTer23 | FS | 0 | LP |
| chr7_150948446_T_TGTCCG | KCNH2 | ENST00000262186 c.2689_2690insCGGAC | ENSP00000262186 p.Lys897ThrfsTer79 | FS | 0 | LP |
| chr7_150950216_G_A | KCNH2 | ENST00000262186 c.2350C>T | ENSP00000262186 p.Arg784Trp | MIS | 0 | LP |
| chr7_150950311_C_T | KCNH2 | ENST00000262186 c.2255G>A | ENSP00000262186 p.Arg752Gln | MIS | 1 | P/LP |
| chr7_150950312_G_A | KCNH2 | ENST00000262186 c.2254C>T | ENSP00000262186 p.Arg752Trp | MIS | 1 | P |
| chr7_150952553_TG_T | KCNH2 | ENST00000262186 c.1428del | ENSP00000262186 p.Asn477MetfsTer44 | FS | 0 | LP |
| chr7_150957291_C_T | KCNH2 | ENST00000262186 c.1128G>A | ENSP00000262186 p.Gln376%3D | SPL | 1 | P/LP |
| chr7_150958128_A_AGGCGC | KCNH2 | ENST00000262186 c.846_847insGCGCC | ENSP00000262186 p.Ser283AlafsTer79 | FS | 0 | LP |
| chr7_150958449_G_A | KCNH2 | ENST00000262186  c.526C>T | ENSP00000262186 p.Arg176Trp | MIS | 0 | LP |
| chr7_150974846_C_T | KCNH2 | ENST00000262186  c.172G>A | ENSP00000262186 p.Glu58Lys | MIS | 1 | LP |
| chr10_110644569_C_T | RBM20 | ENST00000369519  c.115C>T | ENSP00000358532 p.Arg39Ter | SG | 0 | P |
| chr10_110820081_GAGGAAC_G | RBM20 | ENST00000369519 c.2565_2570del | ENSP00000358532 p.Gln856_Glu857del | OTH | 0 | LP |
| chr10_110821364_GGAA_G | RBM20 | ENST00000369519 c.2754_2756del | ENSP00000358532 p.Glu918del | OTH | 0 | LP |
| chr10_110821761_TC_T | RBM20 | ENST00000369519 c.3146del | ENSP00000358532 p.Pro1049LeufsTer55 | FS | 0 | P |
| chr10_110831087_G_A | RBM20 | ENST00000369519 c.3478G>A | ENSP00000358532 p.Gly1160Ser | MIS | 0 | LP |
| chr10_119651695_C_A | BAG3 | ENST00000369085  c.20C>A | ENSP00000358081 p.Ser7Ter | SG | 0 | LP |
| chr10_119651767_  ACCCGCAGACCGGCTGG_A | BAG3 | ENST00000369085 c.96_111del | ENSP00000358081 p.Gln33SerfsTer173 | FS | 0 | LP |
| chr10_119677202_C_T | BAG3 | ENST00000369085 c.1648C>T | ENSP00000358081 p.Gln550Ter | SG | 0 | LP |
| chr11_2445128_CGA_C | KCNQ1 | ENST00000155840 c.34_35del | ENSP00000155840 p.Arg12GlufsTer272 | FS | 0 | LP |
| chr11_2528019_G_A | KCNQ1 | ENST00000155840 c.477+1G>A | NA | SPL | 1 | P |
| chr11_2528023_G_A | KCNQ1 | ENST00000155840 c.477+5G>A | NA | SPL | 1 | P |
| chr11_2570652_G_A | KCNQ1 | ENST00000155840  c.502G>A | ENSP00000155840 p.Gly168Arg | MIS | 1 | P |
| chr11_2570670_C_T | KCNQ1 | ENST00000155840  c.520C>T | ENSP00000155840 p.Arg174Cys | MIS | 1 | P/LP |
| chr11_2570713_G_A | KCNQ1 | ENST00000155840  c.563G>A | ENSP00000155840 p.Trp188Ter | SG | 0 | LP |
| chr11_2570719_GGCTGC_G | KCNQ1 | ENST00000155840 c.573_577del | ENSP00000155840 p.Arg192CysfsTer91 | FS | 1 | P |
| chr11_2570724_C_T | KCNQ1 | ENST00000155840  c.574C>T | ENSP00000155840 p.Arg192Cys | MIS | 0 | LP |
| chr11_2570725_G_A | KCNQ1 | ENST00000155840  c.575G>A | ENSP00000155840 p.Arg192His | MIS | 0 | LP |
| chr11_2570740_C_T | KCNQ1 | ENST00000155840  c.590C>T | ENSP00000155840 p.Pro197Leu | MIS | 0 | LP |
| chr11_2572858_AC_A | KCNQ1 | ENST00000155840  c.796del | ENSP00000155840 p.Leu266CysfsTer23 | FS | 1 | P |
| chr11_2572862_T_C | KCNQ1 | ENST00000155840  c.797T>C | ENSP00000155840 p.Leu266Pro | MIS | 1 | P/LP |
| chr11_2585264_A_G | KCNQ1 | ENST00000155840 c.1085A>G | ENSP00000155840 p.Lys362Arg | MIS | 0 | LP |
| chr11_2587630_C_T | KCNQ1 | ENST00000155840 c.1189C>T | ENSP00000155840 p.Arg397Trp | MIS | 0 | LP |
| chr11_2588718_GA_G | KCNQ1 | ENST00000155840 c.1265del | ENSP00000155840 p.Lys422SerfsTer10 | FS | 1 | P |
| chr11_2768881_C_T | KCNQ1 | ENST00000155840 c.1552C>T | ENSP00000155840 p.Arg518Ter | SG | 1 | P |
| chr11_2768917_C_T | KCNQ1 | ENST00000155840 c.1588C>T | ENSP00000155840 p.Gln530Ter | SG | 1 | P |
| chr11_2776055_G_A | KCNQ1 | ENST00000155840 c.1685+1G>A | NA | SPL | 1 | P/LP |
| chr11_2847874_C_CG | KCNQ1 | ENST00000155840 c.1906dup | ENSP00000155840 p.Ala636GlyfsTer16 | FS | 0 | P |
| chr11_2847958_C_G | KCNQ1 | ENST00000155840 c.1986C>G | ENSP00000155840 p.Tyr662Ter | SG | 0 | P |
| chr11_19182744_A_C | CSRP3 | ENST00000265968  c.511T>G | ENSP00000265968 p.Cys171Gly | MIS | 0 | LP |
| chr11_19188286_A_G | CSRP3 | ENST00000265968  c.131T>C | ENSP00000265968 p.Leu44Pro | MIS | 0 | LP |
| chr11_47332189_G_A | MYBPC3 | ENST00000545968 c.3697C>T | ENSP00000442795 p.Gln1233Ter | SG | 1 | P/LP |
| chr11_47332244_C_T | MYBPC3 | ENST00000545968 c.3642G>A | ENSP00000442795 p.Trp1214Ter | SG | 1 | P/LP |
| chr11_47332565_C_T | MYBPC3 | ENST00000545968 c.3627+1G>A | NA | SPL | 1 | P |
| chr11_47332892_G_A | MYBPC3 | ENST00000545968 c.3412C>T | ENSP00000442795 p.Arg1138Cys | MIS | 0 | LP |
| chr11_47333189_C_G | MYBPC3 | ENST00000545968 c.3330+5G>C | NA | SPL | 1 | P/LP |
| chr11_47333192_A_C | MYBPC3 | ENST00000545968 c.3330+2T>G | NA | SPL | 1 | P |
| chr11_47335894_TCCACGCTGTAG_T | MYBPC3 | ENST00000545968 c.2709_2719del | ENSP00000442795 p.Tyr904ValfsTer143 | FS | 1 | P |
| chr11_47337539_C_T | MYBPC3 | ENST00000545968 c.2454G>A | ENSP00000442795 p.Trp818Ter | SG | 1 | P |
| chr11_47337729_A_AC | MYBPC3 | ENST00000545968 c.2373_2374insG | ENSP00000442795 p.Trp792ValfsTer41 | FS | 1 | P |
| chr11_47338519_C_T | MYBPC3 | ENST00000545968 c.2308+1G>A | NA | SPL | 1 | P |
| chr11_47342578_C_G | MYBPC3 | ENST00000545968 c.1624G>C | ENSP00000442795 p.Glu542Gln | MIS | 1 | P |
| chr11_47342611_C_G | MYBPC3 | ENST00000545968 c.1591G>C | ENSP00000442795 p.Gly531Arg | MIS | 0 | LP |
| chr11_47342698_G_A | MYBPC3 | ENST00000545968 c.1504C>T | ENSP00000442795 p.Arg502Trp | MIS | 1 | P/LP |
| chr11_47342718_C_T | MYBPC3 | ENST00000545968 c.1484G>A | ENSP00000442795 p.Arg495Gln | MIS | 1 | P/LP |
| chr11_47342734_C_T | MYBPC3 | ENST00000545968 c.1468G>A | ENSP00000442795 p.Gly490Arg | MIS | 0 | LP |
| chr11_47343342_C_T | MYBPC3 | ENST00000545968  c.1224-80G>A | NA | OTH | 1 | LP |
| chr11_47346365_G_T | MYBPC3 | ENST00000545968  c.932C>A | ENSP00000442795 p.Ser311Ter | SG | 1 | P |
| chr11_47346372_T_C | MYBPC3 | ENST00000545968  c.927-2A>G | NA | SPL | 1 | P |
| chr11_47346379_C_T | MYBPC3 | ENST00000545968 c  .927-9G>A | NA | SPL | 1 | P |
| chr11_47348424_C_T | MYBPC3 | ENST00000545968  c.772G>A | ENSP00000442795 p.Glu258Lys | MIS | 1 | P/LP |
| chr11_47350501_C_T | MYBPC3 | ENST00000545968 c.406+1G>A | NA | SPL | 1 | LP |
| chr11_47351343_CGTGTGCCCTCT_C | MYBPC3 | ENST00000545968 c.177_187del | ENSP00000442795 p.Glu60AlafsTer49 | FS | 1 | P/LP |
| chr11_47351507_T_C | MYBPC3 | ENST00000545968  c.26-2A>G | NA | SPL | 1 | P/LP |
| chr12_2593255_G_A | CACNA1C | ENST00000399655 c.2573G>A | ENSP00000382563 p.Arg858His | MIS | 1 | P/LP |
| chr12_2607117_G_A | CACNA1C | ENST00000399655 c.3343G>A | ENSP00000382563 p.Glu1115Lys | MIS | 0 | LP |
| chr12_32792703_CA_C | PKP2 | ENST00000340811 c.2385del | ENSP00000342800 p.Ala796LeufsTer91 | FS | 0 | LP |
| chr12_32802499_GGGTGT_G | PKP2 | ENST00000340811 c.2066_2070del | ENSP00000342800 p.His689ProfsTer8 | FS | 1 | P |
| chr12_32821439_A_G | PKP2 | ENST00000340811 c.1930T>C | ENSP00000342800 p.Ser644Pro | MIS | 0 | LP |
| chr12_32824047_TG_T | PKP2 | ENST00000340811 c.1671del | ENSP00000342800 p.Asp557GlufsTer55 | FS | 1 | P |
| chr12_32841027_C_T | PKP2 | ENST00000340811 c.1556+1G>A | NA | SPL | 1 | P/LP |
| chr12_32850891_GC_G | PKP2 | ENST00000340811 c.1252del | ENSP00000342800 p.Ala418ProfsTer2 | FS | 1 | P/LP |
| chr12_32850907_G_A | PKP2 | ENST00000340811 c.1237C>T | ENSP00000342800 p.Arg413Ter | SG | 1 | P |
| chr12_32869034_G_A | PKP2 | ENST00000340811 c.1063C>T | ENSP00000342800 p.Arg355Ter | SG | 1 | P/LP |
| chr12_32877908_C_CAG | PKP2 | ENST00000340811 c.971_972insCT | ENSP00000342800 p.Ala325TrpfsTer28 | FS | 0 | LP |
| chr12_32877910_CCT_C | PKP2 | ENST00000340811 c.968_969del | ENSP00000342800 p.Gln323ArgfsTer12 | FS | 0 | LP |
| chr12_32878041_ACG_A | PKP2 | ENST00000340811 c.837_838del | ENSP00000342800 p.Val280HisfsTer55 | FS | 1 | P/LP |
| chr12_32878512_C_T | PKP2 | ENST00000340811  c.368G>A | ENSP00000342800 p.Trp123Ter | SG | 1 | P |
| chr12_32896566_  TCCGCAGGCTCTTGACTGTCTGG_T | PKP2 | ENST00000340811 c.144_165del | ENSP00000342800 p.Gln49SerfsTer56 | FS | 1 | P/LP |
| chr12_32896580_ACTGT_A | PKP2 | ENST00000340811 c.148_151del | ENSP00000342800 p.Thr50SerfsTer61 | FS | 1 | P |
| chr12_110911165_A_T | MYL2 | ENST00000228841  c.413T>A | ENSP00000228841 p.Met138Lys | MIS | 0 | LP |
| chr12_110913097_T_G | MYL2 | ENST00000228841  c.401A>C | ENSP00000228841 p.Glu134Ala | MIS | 0 | LP |
| chr12_110914276_T_A | MYL2 | ENST00000228841  c.184A>T | ENSP00000228841 p.Lys62Ter | SG | 0 | P |
| chr12_110915787_A_G | MYL2 | ENST00000228841  c.97T>C | ENSP00000228841 p.Phe33Leu | MIS | 0 | LP |
| chr12_110919149_GT_G | MYL2 | ENST00000228841  c.47del | ENSP00000228841 p.Asn16ThrfsTer34 | FS | 0 | LP |
| chr14_23388250_G_A | MYH6 | ENST00000405093 c.4264C>T | ENSP00000386041 p.Arg1422Trp | MIS | 0 | LP |
| chr14_23415225_C_T | MYH7 | ENST00000355349 c.5329G>A | ENSP00000347507 p.Ala1777Thr | MIS | 0 | LP |
| chr14_23415228_T_C | MYH7 | ENST00000355349 c.5326A>G | ENSP00000347507 p.Ser1776Gly | MIS | 0 | LP |
| chr14_23415267_C_T | MYH7 | ENST00000355349 c.5287G>A | ENSP00000347507 p.Ala1763Thr | MIS | 0 | LP |
| chr14_23415651_C_T | MYH7 | ENST00000355349 c.5135G>A | ENSP00000347507 p.Arg1712Gln | MIS | 1 | P |
| chr14_23416980_T_A | MYH7 | ENST00000355349 c.4532A>T | ENSP00000347507 p.Asp1511Val | MIS | 0 | LP |
| chr14_23417174_G_A | MYH7 | ENST00000355349 c.4498C>T | ENSP00000347507 p.Arg1500Trp | MIS | 1 | P/LP |
| chr14_23422256_C_T | MYH7 | ENST00000355349 c.3169G>A | ENSP00000347507 p.Gly1057Ser | MIS | 1 | LP |
| chr14_23422292_G_A | MYH7 | ENST00000355349 c.3133C>T | ENSP00000347507 p.Arg1045Cys | MIS | 0 | LP |
| chr14_23424047_C_T | MYH7 | ENST00000355349 c.2782G>A | ENSP00000347507 p.Asp928Asn | MIS | 1 | P/LP |
| chr14_23424107_G_C | MYH7 | ENST00000355349 c.2722C>G | ENSP00000347507 p.Leu908Val | MIS | 1 | P |
| chr14_23424112_T_C | MYH7 | ENST00000355349 c.2717A>G | ENSP00000347507 p.Asp906Gly | MIS | 1 | P |
| chr14_23424839_C_T | MYH7 | ENST00000355349 c.2609G>A | ENSP00000347507 p.Arg870His | MIS | 1 | P |
| chr14_23424842_C_T | MYH7 | ENST00000355349 c.2606G>A | ENSP00000347507 p.Arg869His | MIS | 1 | LP |
| chr14_23427657_C_T | MYH7 | ENST00000355349 c.1816G>A | ENSP00000347507 p.Val606Met | MIS | 1 | P/LP |
| chr14_23427746_T_C | MYH7 | ENST00000355349 c.1727A>G | ENSP00000347507 p.His576Arg | MIS | 1 | P/LP |
| chr14_23428587_C_A | MYH7 | ENST00000355349 c.1491G>T | ENSP00000347507 p.Glu497Asp | MIS | 1 | P/LP |
| chr14_23428642_T_C | MYH7 | ENST00000355349 c.1436A>G | ENSP00000347507 p.Asn479Ser | MIS | 1 | P/LP |
| chr14_23428992_A_G | MYH7 | ENST00000355349 c.1370T>C | ENSP00000347507 p.Ile457Thr | MIS | 1 | LP |
| chr14_23429038_G_A | MYH7 | ENST00000355349 c.1324C>T | ENSP00000347507 p.Arg442Cys | MIS | 1 | P/LP |
| chr14_23430601_C_T | MYH7 | ENST00000355349  c.958G>A | ENSP00000347507 p.Val320Met | MIS | 1 | P/LP |
| chr14_23433086_A_G | MYH7 | ENST00000355349  c.343T>C | ENSP00000347507 p.Tyr115His | MIS | 0 | LP |
| chr15_34790434_A_G | ACTC1 | ENST00000290378 c.1112T>C | ENSP00000290378 p.Ile371Thr | MIS | 0 | LP |
| chr15_34790458_T_C | ACTC1 | ENST00000290378 c.1088A>G | ENSP00000290378 p.Glu363Gly | MIS | 0 | LP |
| chr15_34792089_C_G | ACTC1 | ENST00000290378 c.808+1G>C | NA | SPL | 0 | LP |
| chr15_34793420_G_T | ACTC1 | ENST00000290378  c.279C>A | ENSP00000290378 p.Tyr93Ter | SG | 0 | LP |
| chr15_63044030_G_T | TPM1 | ENST00000403994  c.118G>T | ENSP00000385107 p.Glu40Ter | SG | 0 | LP |
| chr17_39665425_G_A | TCAP | ENST00000309889  c.66G>A | ENSP00000312624 p.Trp22Ter | SG | 1 | P/LP |
| chr17_70175239_G_A | KCNJ2 | ENST00000243457  c.200G>A | ENSP00000243457 p.Arg67Gln | MIS | 1 | P/LP |
| chr18_31068135_T_TCTTC | DSC2 | ENST00000280904 c.2585_2586insGAAG | ENSP00000280904 p.Gly863LysfsTer13 | FS | 0 | LP |
| chr18_31071603_AC_A | DSC2 | ENST00000280904 c.2125+1del | NA | SPL | 0 | LP |
| chr18_31071797_A_AG | DSC2 | ENST00000280904 c.1932_1933insC | ENSP00000280904 p.Ser645LeufsTer11 | FS | 0 | LP |
| chr18_31074713_G_A | DSC2 | ENST00000280904 c.1858C>T | ENSP00000280904 p.Gln620Ter | SG | 1 | P |
| chr18_31083061_C_T | DSC2 | ENST00000280904  c.943-1G>A | NA | SPL | 0 | LP |
| chr18_31101936_TCC_T | DSC2 | ENST00000280904 c.34_35del | ENSP00000280904 p.Gly12SerfsTer18 | FS | 0 | LP |
| chr18_31521136_TAG_T | DSG2 | ENST00000261590 c.420_421del | ENSP00000261590 p.Lys141ThrfsTer7 | FS | 0 | LP |
| chr18_31521244_G_C | DSG2 | ENST00000261590 c.523+1G>C | NA | SPL | 1 | P/LP |
| chr18_31522250_G_A | DSG2 | ENST00000261590 c.690+1G>A | NA | SPL | 0 | LP |
| chr18_31524582_AGTG_A | DSG2 | ENST00000261590 c.828_828+2del | NA | SPL | 0 | P |
| chr18_31541284_GA_G | DSG2 | ENST00000261590 c.1973del | ENSP00000261590 p.Asn658IlefsTer24 | FS | 0 | LP |
| chr18_31542627_CA_C | DSG2 | ENST00000261590 c.2110del | ENSP00000261590 p.Ile704LeufsTer37 | FS | 0 | LP |
| chr18_31546258_CCA_C | DSG2 | ENST00000261590 c.2875_2876del | ENSP00000261590 p.Gln959GlufsTer22 | FS | 0 | P |
| chr18_31546440_AAGAG_A | DSG2 | ENST00000261590 c.3059_3062del | ENSP00000261590 p.Glu1020AlafsTer18 | FS | 0 | P |
| chr18_31546440_AAGAG_AAG | DSG2 | ENST00000261590 c.3061_3062del | ENSP00000261590 p.Ser1021LeufsTer16 | FS | 0 | LP |
| chr18_31546528_GAA_G | DSG2 | ENST00000261590 c.3144_3145del | ENSP00000261590 p.Arg1049SerfsTer17 | FS | 0 | LP |
| chr18_31592974_G_A | TTR | ENST00000237014  c.148G>A | ENSP00000237014 p.Val50Met | MIS | 1 | P |
| chr18_31595152_T_A | TTR | ENST00000237014  c.233T>A | ENSP00000237014 p.Leu78His | MIS | 1 | P/LP |
| chr19_55151856_C_T | TNNI3 | ENST00000344887  c.611G>A | ENSP00000341838 p.Arg204His | MIS | 1 | P/LP |
| chr19_55154094_C_T | TNNI3 | ENST00000344887  c.485G>A | ENSP00000341838 p.Arg162Gln | MIS | 1 | P/LP |
| chr19_55154145_C_T | TNNI3 | ENST00000344887  c.434G>A | ENSP00000341838 p.Arg145Gln | MIS | 1 | P/LP |
| chr19_55156205_A_G | TNNI3 | ENST00000344887  c.278T>C | ENSP00000341838 p.Leu93Pro | MIS | 0 | LP |

**Footnotes/Abbreviations**: 1: Variants are described above as chromosome_position_reference_alternate, with position provided per GRCh38; 3: FS=frameshift, SG=stop gain, MIS=missense; 4: indicates that variant was classified as LP/P based on classification in ClinVar; 5: LP=likely pathogenic, P=pathogenic.

**Supplementary Table 5: Comparison of Variant Consequences Between Known and Novel Variants**

| **Variant Type** | **Variant Consequence** | | | |
| --- | --- | --- | --- | --- |
|  | **Frameshift/Stop** | **Missense** | **Canonical Splice** | **Other** |
| Known (n=136) | 66 (49%) | 43 (32%) | 18 (13%) | 9 (7%) |
| Novel (n=164) | 100 (61%) | 49 (30%) | 13 (8%) | 2 (1%) |
| Total (n=300) | 166 (55%) | 92 (31%) | 31 (10%) | 11 (4%) |

**Supplementary Table 6: Case-Control Analysis for Genes Associated with SCA, Sudden Cardiac Arrest-Total Cohort vs. Control Cohort**

| **Gene** | **CLR-SKAT**  **p-value** |
| --- | --- |
| ACTC1 | 0.287 |
| BAG3 | 0.389 |
| CACNA1C | 0.017 |
| CASQ2 | 0.561 |
| CSRP3 | 0.783 |
| DES | 0.507 |
| DSC2 | 0.238 |
| DSG2 | 0.008 |
| DSP | 0.405 |
| EYA4 | 0.621 |
| FLNC | 0.208 |
| KCNH2 | 0.882 |
| KCNJ2 | 0.621 |
| KCNQ1 | 0.747 |
| LMNA | 0.008 |
| MYBPC3 | 0.269 |
| MYH6 | 0.429 |
| MYH7 | 0.256 |
| MYL2 | 1.000 |
| MYL3 | 0.243 |
| NEXN | 0.580 |
| PKP2 | 0.649 |
| RBM20 | 0.201 |
| RYR2 | 0.833 |
| SCN5A | 1.000 |
| SGCD | 0.448 |
| TCAP | 0.243 |
| TNNI3 | 0.287 |
| TNNT2 | 0.071 |
| TPM1 | 0.429 |
| TTN | 0.740 |
| TTR | 0.359 |

**Supplementary Table 7: Case-Control Analysis for Genes Associated with SCA, Sudden Cardiac Arrest-Nonischemic Subgroup Matched 1:6 to Control Cohort**

| **Gene** | **CLR-SKAT**  **p-value** |
| --- | --- |
| ACTC1 | 0.677 |
| BAG3 | 0.047 |
| CASQ2 | 0.047 |
| CSRP3 | 0.677 |
| DES | 0.386 |
| DSC2 | 0.047 |
| DSG2 | 0.845 |
| DSP | 0.495 |
| EYA4 | 0.677 |
| FLNC | 0.007 |
| KCNH2 | 0.034 |
| KCNQ1 | 0.039 |
| LMNA | 0.003 |
| MYBPC3 | 0.019 |
| MYH6 | 0.677 |
| MYH7 | 0.138 |
| MYL2 | 0.362 |
| MYL3 | 0.677 |
| NEXN | 0.677 |
| PKP2 | 0.050 |
| RBM20 | 0.719 |
| RYR2 | 0.392 |
| SCN5A | 0.835 |
| SGCD | 0.845 |
| TCAP | 0.677 |
| TNNI3 | 0.014 |
| TNNT2 | 0.677 |
| TPM1 | 0.677 |
| TTN | 0.022 |
| TTR | 0.845 |

**Supplementary Table 8: Case-Control Analysis for Genes Associated with SCA, Sudden Cardiac Arrest-Ischemic Subgroup Matched 1:6 to Control Cohort**

| **Gene** | **CLR-SKAT**  **p-value** |
| --- | --- |
| ACTC1 | 0.137 |
| CACNA1C | 0.003 |
| CASQ2 | 0.916 |
| CSRP3 | 0.845 |
| DES | 0.361 |
| DSC2 | 0.275 |
| DSG2 | 5.20 x 10^-5^ |
| DSP | 0.274 |
| EYA4 | 0.677 |
| FLNC | 0.022 |
| KCNH2 | 0.998 |
| KCNQ1 | 0.591 |
| LMNA | 0.014 |
| MYBPC3 | 0.309 |
| MYH6 | 0.527 |
| MYH7 | 0.063 |
| MYL2 | 1.000 |
| MYL3 | 0.386 |
| NEXN | 0.386 |
| PKP2 | 0.480 |
| RBM20 | 0.092 |
| RYR2 | 0.886 |
| SCN5A | 0.625 |
| SGCD | 0.099 |
| TCAP | 0.386 |
| TNNI3 | 0.916 |
| TNNT2 | 4.09 x 10^-4^ |
| TPM1 | 0.527 |
| TTN | 0.767 |
| TTR | 0.334 |

**Supplementary Table 9: Case-Control Analysis for Genes Associated with SCA, Sudden Cardiac Arrest-Unassigned Subgroup Matched 1:6 to Control Cohort**

| **Gene** | **CLR-SKAT**  **p-value** |
| --- | --- |
| ACTC1 | 0.677 |
| CASQ2 | 0.677 |
| CSRP3 | 0.677 |
| DES | 0.099 |
| DSG2 | 0.034 |
| DSP | 0.680 |
| EYA4 | 0.677 |
| FLNC | 0.095 |
| KCNH2 | 0.219 |
| KCNQ1 | 0.274 |
| MYBPC3 | 0.027 |
| MYH6 | 0.677 |
| MYH7 | 0.812 |
| MYL2 | 0.254 |
| MYL3 | 0.677 |
| NEXN | 0.114 |
| PKP2 | 0.335 |
| RBM20 | 0.719 |
| RYR2 | 0.970 |
| SCN5A | 0.290 |
| SGCD | 0.677 |
| TCAP | 0.677 |
| TNNI3 | 0.845 |
| TPM1 | 0.527 |
| TTN | 0.756 |
| TTR | 0.719 |
